# Hospitalization Patterns, Outcomes, and Resource Utilization among Individuals with Gender Dysphoria: A Nationwide Study

**DOI:** 10.1101/2023.08.25.23294637

**Authors:** Fidelis Uwumiro, Micheal Bojerenu, Chinwendu Obijuru, Okunlola Oluwasemilore, Vivian Nwike, Summayyah Abdul, Halleluya Yeyehyirad, Udegbe Silvia, Olawale Abesin, George Orjih

## Abstract

**Background:** The significance of providing gender-affirming care has gained prominence in recent years. This study examines the hospitalization patterns of individuals with gender dysphoria, exploring the indications for hospitalization, types of gender-affirming care offered, resource utilization, and outcomes.

**Methods:** We analyzed 67,454 discharge records from the 2016-2020 Nationwide Inpatient Sample (NIS). We identified cases of gender dysphoria using the International Classification of Diseases, 10th Revision (ICD-10) code F64.0. Demographic data, co-morbidity scores, indications for hospitalization, care patterns, length of hospital stay, mortality rates, and hospital charges were rigorously examined using ICD-10 codes and survey techniques. The trend in hospital expenditures was assessed using the Cochran-Armitage test with statistical significance set at P<.05.

**Results:** About 53.6% were assigned female at birth, 46.4% male. The mean age was 33 ± 0.2 years. White Americans constituted 67.3%, Blacks 14.9%, Hispanics 10%, and Asians/Pacific Islanders 2.1%. Most admissions (51.6%) were ages 16-35, with 9.5% aged 60 or older. Mental health services (55.2%) and cross-sex hormonal therapy (48.8%) were common. Gender-affirming surgeries were performed in 7.1% of cases, while 0.4% received puberty-suppressing therapy. About 355 mortalities were recorded. Mortality rates increased with advancing age, with the highest mortality among individuals over 60 years old (205). No deaths occurred among those who received gender-affirming surgery. The mean length of hospital stay (LOS) was 6 days. Patients in the age range of 5 to 15 years had longer hospital stays compared to other age groups, with a mean LOS of 7.3 ± 0.2 days (P<0.001). Total hospital costs for gender-affirming care exceeded $3.3 billion, with a notable trend of increasing costs over the study period. The average costs for gender-affirming surgery, mental health services, and cross-sex hormone therapy were $101,349 ± $5,881, $41,938 ± $1,350, and $70,873 ± $13,528, respectively. Over time, mean and total costs for gender dysphoria hospitalizations increased from 400 million in 2016 to 1.3 billion in 2020 (*P*_*trend*_<0.001).

**Conclusion:** This study reinforces the critical role of mental health services and cross-sex hormonal therapy in gender transition. It also challenges stereotypes by demonstrating that gender-affirming surgeries are sought by individuals across a wide age spectrum. However, it underscores the importance of addressing healthcare disparities, particularly for older individuals. The substantial financial costs associated with gender-affirming care underscore its significance and increasing demand. These findings emphasize the evolving healthcare needs of the gender dysphoria population, necessitating ongoing research and comprehensive support to advance gender-affirming care and understanding.

## INTRODUCTION

Transgender and gender non-conforming (TGNC) individuals who experience a misalignment between their gender identity and assigned sex at birth represent a diverse population with unique and individualized healthcare needs. In recent years, the visibility and recognition of TGNC individuals have grown significantly [1-2]. Alongside this progress, the importance of providing gender-affirming care has gained prominence in the healthcare landscape [3]. Gender-affirming care includes a range of medical interventions tailored to support the physical, emotional, and psychological well-being of gender dysphoria individuals [4,5].

Transgender and gender non-conforming are inclusive terms that encompass individuals whose gender identity or expression differs from the sex assigned to them at birth. This includes people who identify as transgender, non-binary, genderqueer, genderfluid or any other gender identity beyond the traditional male and female binary. On the other hand, gender dysphoria is a clinical manifestation of significant distress, discomfort, and/or depressive states, secondary to the incongruence of one’s gender identity and the sex assigned at birth, markedly affecting one’s well-being [6]. It is recognized as a medical diagnosis in both the DSM-5 (Diagnostic and Statistical Manual of Mental Disorders) and ICD (International Classification of Diseases) [7]. The coding for gender dysphoria underwent significant changes between ICD-10 and ICD-11. In ICD-10, it was classified as “F64.0,” a psychiatric disorder. However, ICD-11 shifted to “Conditions Related to Sexual Health” with the code “HA60.0,” acknowledging gender diversity as a natural human variation, not a mental illness. This reclassification represents a significant step in destigmatizing and affirming the experiences of transgender and gender non-conforming individuals in healthcare settings.

Understanding hospitalization patterns among gender dysphoria individuals is useful for developing tailored interventions and practices that promote equitable and inclusive healthcare services for this vulnerable population. Studies have indicated that transgender individuals often face unique healthcare challenges that can impact their likelihood of hospitalization. Factors such as discrimination, lack of culturally competent care, and socioeconomic disparities contribute to the disparities in healthcare access and utilization for this population [8-12]. Research has shown that transgender individuals may experience higher rates of hospitalization for certain conditions, including mental health issues, gender-affirming surgeries, and complications related to hormone therapy [13]. Despite these important initial findings, little is known about the hospitalization patterns, outcomes, and resource utilization of gender dysphoria individuals in the United States.

Our primary goal is to explore the hospitalization patterns among individuals with gender dysphoria. To achieve this, we aim to examine various aspects, including the main reasons for hospitalization, the types of gender-affirming care often provided, the resource utilization (hospital stays and the total healthcare costs), and hospitalization outcomes associated with gender dysphoria.

## MATERIALS AND METHODS

A total of 35 million discharge records from the 2016-2020 Nationwide Inpatient Sample (NIS) were obtained from the Agency for Healthcare Research and Quality’s Healthcare Cost and Utilization Project. NIS is the largest publicly available all-payer database, covering patients from Medicare, Medicaid, private insurance, and the uninsured. The database rigorously reflects a 20% sample of all U.S. hospital admissions, excluding admissions to rehabilitation and federal hospitals (e.g., Veterans Affairs hospitals). The 2020 NIS sample includes data from all participating states (46 states and the District of Columbia), covering 98% of the U.S. population. Each year’s NIS data contains around 7-8 million records, each with a corresponding discharge weight. The representative sampling and tabulation of discharge weights are achieved by characterizing participating institutions across 5 strata: ownership or control, bed size, teaching status, urban or rural location, and US region. Weighted, each year’s NIS data estimates approximately 36 million hospital stays [14].

The International Classification of Diseases, 10th Revision (ICD-10), was used to identify discharge records with the gender dysphoria diagnostic code of F64.0. All discharges for individuals 5 years or older were included for analysis. Discharge records with missing or incomplete data were excluded from the analysis. The study variables included demographic information of patients and hospitals including age, gender, race, median annual income, hospital region, bed size, and teaching status among others. These details were already predefined and available within the NIS sample. The burden of co-morbidity was assessed using the combined Charlson comorbidity index score. The indications for hospitalization and pattern of care received by gender dysphoria patients were identified and classified using a combination of Major Diagnostic Categories (MDC) based on organ systems and transition-related services such as cross-sex hormone therapy, puberty-suppressing therapy, and gender-affirming surgery (Appendix 1) using ICD-10 diagnostic/procedure codes. Age categories were defined as 5-15, 16-25, 26-35, 36-45, 46-60, and >60 years.

The study aimed to characterize hospitalizations for gender dysphoria patients based on sociodemographics and clinical indications. Secondary outcomes included hospital stay length, gender-affirming care patterns, discharge survival, total hospital costs, and trends in hospitalization over the study period. Stata statistical software v.17.0MP (StataCorp LLC, College Station, TX, USA) and survey techniques were used for estimation, presenting means, frequencies, and proportions. Inflation was adjusted for using the consumer price index. Linear regression was used to compare multigroup variables, Pearson’s χ2 test analyzed categorical variables, and Cochran-Armitage tests were used to examine trends over time. Statistically significant values were denoted by p<0.05.

## RESULTS

### Sociodemographic characteristics

A total of 67,454 discharges were analyzed. Of these admissions, approximately 53.6% were assigned female at birth, while 46.4% were assigned male at birth. The mean age of the study population was 33 ± 0.2 years. The majority of admissions (51.6%) involved individuals aged 16 to 35 years, while 9.5% (6,408) of patients were 60 years or older. Most of the hospitalizations were covered by insurance (94.3%) and were evenly distributed across all quartiles of median annual income, as indicated in Table 1. Urban hospitals accounted for the majority of admissions (95.6%), spanning different regions of the United States. Regarding the racial composition of the study population, White Americans comprised the largest portion, accounting for 67.3% of the total cohort. Blacks represented 14.9% of the population, Hispanics accounted for 10%, and Asians or Pacific Islanders made up 2.1%.

**Table 1:** Sociodemographic characteristics.

| Variable | n, % Unless Otherwise Specified |
| --- | --- |
| Total admissions for patients with gender dysphoria | 67,454 |
| Sex assigned at birth |  |
| Female | 36,155 (53.6) |
| Male | 31,298 (46.4) |
| Age categories, years |  |
| 5 – 15 | 8,904 (13.2) |
| 16 – 25 | 20,439 (30.3) |
| 26 – 35 | 14,368 (21.3) |
| 36 – 45 | 8,161 (12.1) |
| 46 – 60 | 9,241 (13.7) |
| > 60 | 6,408 (9.5) |
| Mean age, years | 33 ± 0.2 |
| Mean hospital charge, USD | 49,830 ± 1,367 |
| Aggregate hospital charge, Billion USD | 3.33 |
| Elective admission | 14,097 (20.9) |
| Emergency admission | 53,356 (79.1) |
| The mean length of stay, days | 14.7 ± 0.1 |
| Discharge disposition |  |
| Died in the hospital | 358 (0.53) |
| Routine home discharge | 54,368 (80.6) |
| Transfer to a short-term hospital | 877 (1.3) |
| Transfer to other skilled care<br>(Including SNF, home health care & ICF) | 10,118 (15.0) |
| Against medical advice | 1,754 (2.6) |
| Discharge quarter |  |
| First quarter | 15,987 (23.7) |
| Second quarter | 15,245 (22.6) |
| Third quarter | 17,201 (25.5) |
| Fourth quarter | 19,022 (28.2) |
| Race |  |
| White American | 45,396 (67.3) |
| Black | 10,051 (14.9) |
| Hispanic | 6,745 (10.0) |
| Asian or Pacific Islander | 1,417 (2.1) |
| Native American | 675 (1.0) |
| Others | 3,238 (4.8) |
| Weekend admission | 12,007 (17.8) |
| Combined Charlson comorbidity index |  |
| 0 | 41,754 (61.9) |
| 1 | 12,614 (18.7) |
| 2 | 4,317 (6.4) |
| ≥3 | 8,769 (13.0) |
| Median annual income quartiles |  |
| 1–43,999 | 18,820 (27.9) |
| 44,000–55,999 | 17,201 (25.5) |
| 56,000–73,999 | 16,391 (24.3) |
| ≥ 74,000 | 15,042 (22.3) |
| Insurance status |  |
| Medicare | 12,142 (18.0) |
| Medicaid | 25,160 (37.3) |
| Private including HMO | 26,375 (39.1) |
| Self-pay | 3,845 (5.7) |
| Hospital location |  |
| Rural | 2,968 (4.4) |
| Urban-non-teaching | 9,713 (14.4) |
| Urban teaching hospital | 54,773 (81.2) |
| Hospital region |  |
| Northeast | 16,054 (23.8) |
| Midwest | 17,066 (25.3) |
| South | 16,391 (24.3) |
| West | 18,010 (26.7) |
| Hospital bed size |  |
| Small | 15,582 (23.1) |
| Medium | 16,054 (23.8) |
| Large | 35,818 (53.1) |

**\*\***Defined by the Major Diagnostic Categories (MDC) codes in use on discharge date
<sup>a</sup> Sundararajan's adaptation of the modified Deyo's Charlson Comorbidity Index (CCI) offers a refined approach for population-based investigations. This adaptation classifies the CCI into four distinct groups, each indicative of escalating mortality risk. A CCI score surpassing 3 is associated with an approximate 25% 10-year mortality rate, whereas scores of 2 or 1 correspond to 10% and 4% 10-year mortality rates, respectively.
USD, United States dollar; HMO, health maintenance organization

### Transition-related services

We examined the care sought by individuals with gender dysphoria, including both transition-related and other general medical care. We categorized all possible diagnoses into 25 distinct areas based on organ systems. Within the study period, approximately 55.2% (37,235) of all hospitalizations for gender dysphoria involved services related to mental diseases and disorders. Cross-sex hormonal therapy was received by around 48.8% of individuals, while 7.1% (4,770) underwent gender-affirming surgery, and 0.4% (260) received puberty-suppressing therapy. Up to 5.7% were hospitalized for injuries, poisoning, and other toxic effects of drugs within the same period. Table 2 summarizes other care received by Individuals with gender dysphoria in the study. Mental health services and cross-sex hormonal therapy emerged as the most commonly provided services across all age groups. The frequency of transition-related services varied depending on the age category (P<0.001; Fig 1).

**Table 2:**
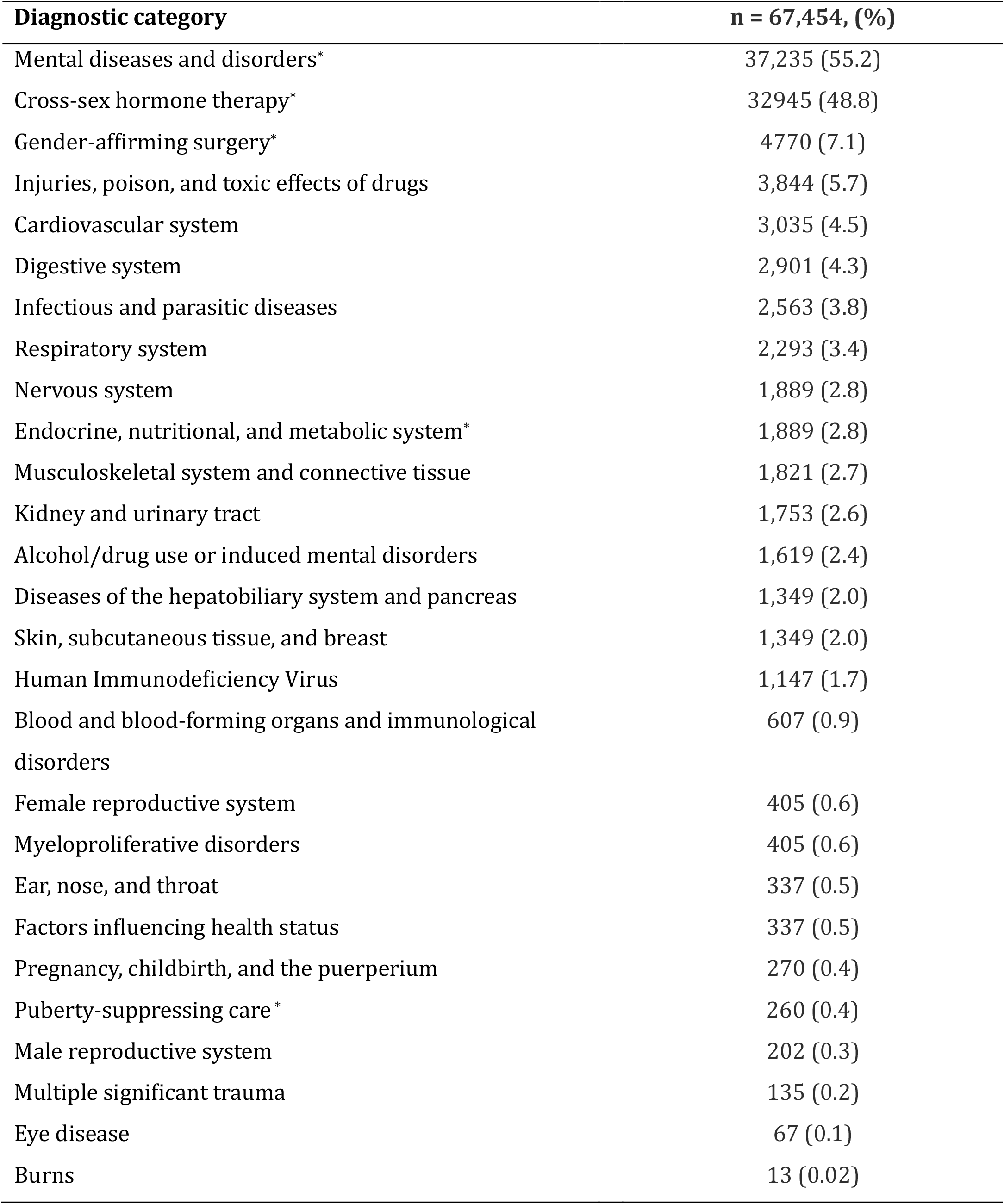
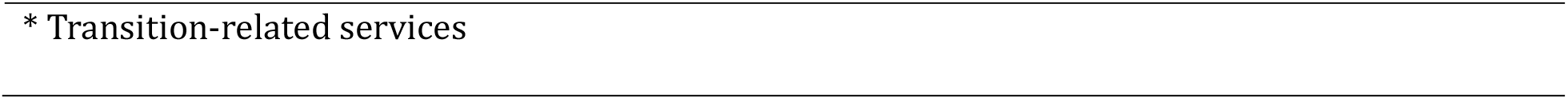
Care received by gender dysphoria individuals: classified by Major Diagnostic Categories (MDC) in use on the discharge day.

| **Diagnostic category** | **n = 67,454, (%)** |
| --- | --- |
| Mental diseases and disorders* | 37,235 (55.2) |
| Cross-sex hormone therapy* | 32945 (48.8) |
| Gender-affirming surgery* | 4770 (7.1) |
| Injuries, poison, and toxic effects of drugs | 3,844 (5.7) |
| Cardiovascular system | 3,035 (4.5) |
| Digestive system | 2,901 (4.3) |
| Infectious and parasitic diseases | 2,563 (3.8) |
| Respiratory system | 2,293 (3.4) |
| Nervous system | 1,889 (2.8) |
| Endocrine, nutritional, and metabolic system* | 1,889 (2.8) |
| Musculoskeletal system and connective tissue | 1,821 (2.7) |
| Kidney and urinary tract | 1,753 (2.6) |
| Alcohol/drug use or induced mental disorders | 1,619 (2.4) |
| Diseases of the hepatobiliary system and pancreas | 1,349 (2.0) |
| Skin, subcutaneous tissue, and breast | 1,349 (2.0) |
| Human Immunodeficiency Virus | 1,147 (1.7) |
| Blood and blood-forming organs and immunological disorders | 607 (0.9) |
| Female reproductive system | 405 (0.6) |
| Myeloproliferative disorders | 405 (0.6) |
| Ear, nose, and throat | 337 (0.5) |
| Factors influencing health status | 337 (0.5) |
| Pregnancy, childbirth, and the puerperium | 270 (0.4) |
| Puberty-suppressing care * | 260 (0.4) |
| Male reproductive system | 202 (0.3) |
| Multiple significant trauma | 135 (0.2) |
| Eye disease | 67 (0.1) |
| Burns | 13 (0.02) |

\* Transition-related services

**Figure 1:**
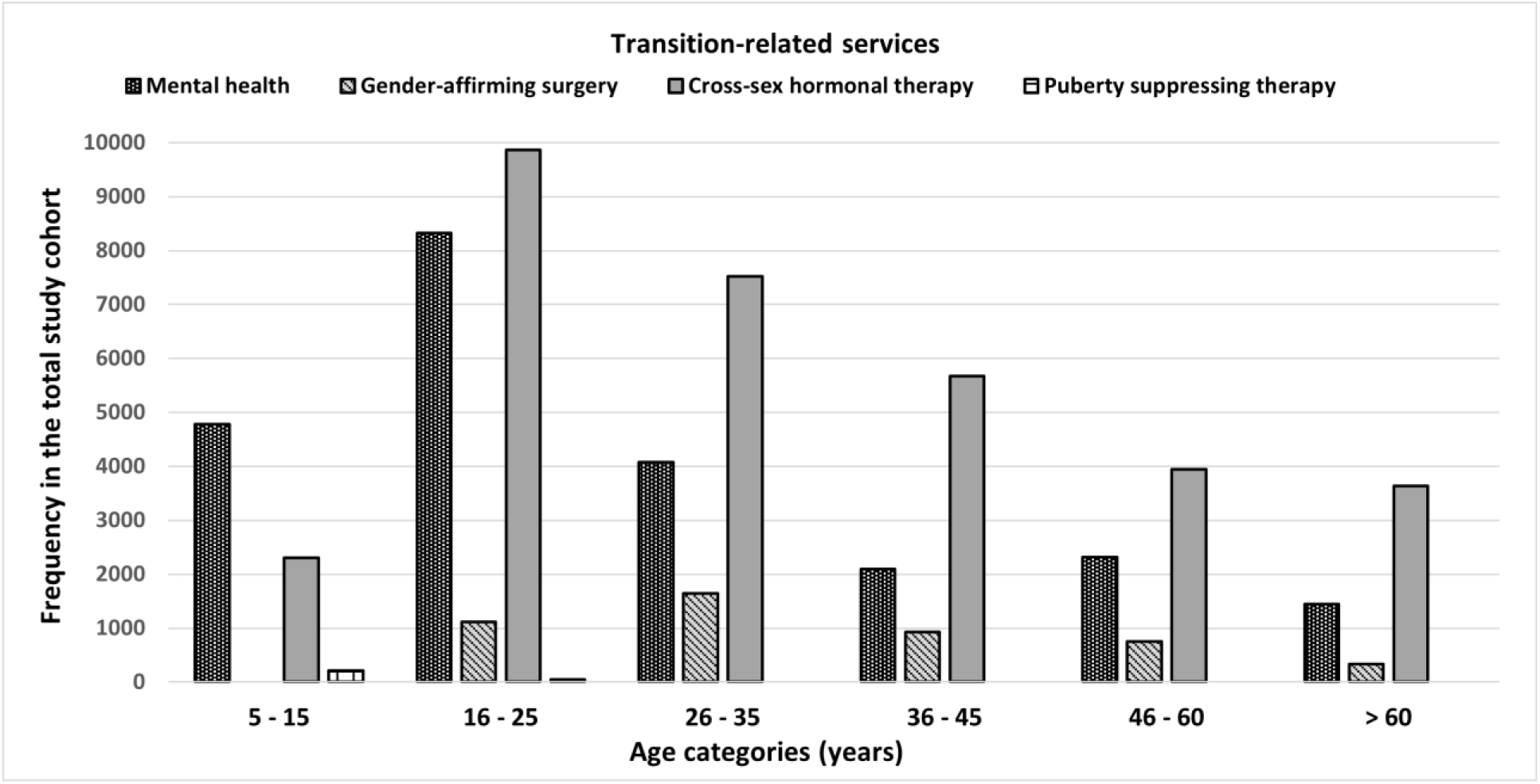
Transition-related care by age categories.

### Gender-affirming surgeries

About 4,770 (7.1%) discharges involved gender-affirming surgeries. Table 3 provides an overview of the different types of gender-affirming surgeries performed on both transmasculine and transfeminine individuals within the study cohort. The majority of surgeries were conducted on individuals between the ages of 26 and 35, followed by those in the age range of 16 to 25. Notably, the data reveals that over 500 surgeries were performed on individuals aged 60 and above (Table 3). Caucasians had the highest number of gender-affirming surgeries followed by Hispanics and Blacks respectively.

**Table 3:** Gender-affirming surgeries performed among gender dysphoria individuals hospitalized between 2016 and 2020: stratified by age category and race.

| **Age Categories/**<br>**Race** | **Male-to-female chest**<br>**surgeries**<br>**(n = 1,430)** | **Male-to-female genital**<br>**surgeries (n = 2,990)** | **Male-to-female facial**<br>**feminization**<br>**surgery**<br>**(n = 620)** | **Female-to-male**<br>**surgeries**<br>**(n = 2,845)** | **Total** |
| --- | --- | --- | --- | --- | --- |
| 5 – 15 | 0 | 0 | 0 | 0 | 0 |
| 16 – 25* | 365 | 640 | 120 | 710 | 1,835 |
| 26 – 35 | 450 | 1,015 | 250 | 950 | 2,665 |
| 36 – 45 | 310 | 615 | 75 | 605 | 1,605 |
| 46 – 60 | 210 | 515 | 100 | 410 | 1,235 |
| > 60 | 95 | 205 | 75 | 170 | 545 |
| Caucasian | 860 | 1,974 | 360 | 1,700 | 4,894 |
| Blacks | 150 | 250 | 95 | 315 | 810 |
| Hispanics | 160 | 300 | 65 | 300 | 825 |
| Asians/Pacific<br>Islanders | 45 | 80 | 35 | 100 | 260 |
| Native<br>Americans | 10 | 10 | 0 | 15 | 35 |
| Other races | 130 | 255 | 50 | 310 | 745 |
| * Includes 45 male-to-female and 10 female-to-male surgeries performed before the age of 18 years |  |  |  |  |  |

### Hospital length of stay, mortality, and total hospital charges

The overall mean length of hospital stay in the study was 6 ± 0.1 days. Interestingly, patients in the age range of 5 to 15 years had longer hospital stays compared to other age groups, with an average of 7.3 ± 0.2 days (P<0.001). Furthermore, individuals who underwent any gender-affirming surgery had a shorter mean length of stay compared to those who did not (6.1 ± 0.1 days vs. 3.7 ± 0.2 days; P=0.03).

Throughout the study period, a total of 355 mortalities were recorded. Among different racial groups, White Americans had the highest number of deaths (205), followed by Blacks (70), Hispanics (59), Asians/Pacific Islanders (5), Native Americans (5), and individuals from other races (5). Mortality rates increased with advancing age, with the highest number of deaths occurring among individuals over 60 years old (205), followed by those in the age range of 46 to 60 years (70), and individuals aged 36 to 45 years (30). Notably, no mortality was recorded among individuals who received gender-affirming surgery. Among those who received mental health services, 54 deaths were recorded.

The total hospital costs for all admissions amounted to 3.33 billion USD, with an average cost of $49,830 ± $1,367 overall. More healthcare expenditure was allocated to other transition-related services and general care compared to gender-affirming procedures, with costs reaching 2.85 billion USD and 481 million USD, respectively (P<0.001).

When examining specific services, the average costs for care related to gender-affirming surgery, mental health services, and cross-sex hormone therapy (including puberty-suppressing therapy) were $101,349 ± $5,881, $41,938 ± $1,350, and $70,873 ± $13,528, respectively. On further trend analysis, the mean and aggregate costs of hospitalization for gender dysphoria individuals were observed to have increased steadily over time (*Ptrend*<0.001; Figure 2).

**Figure 2:**
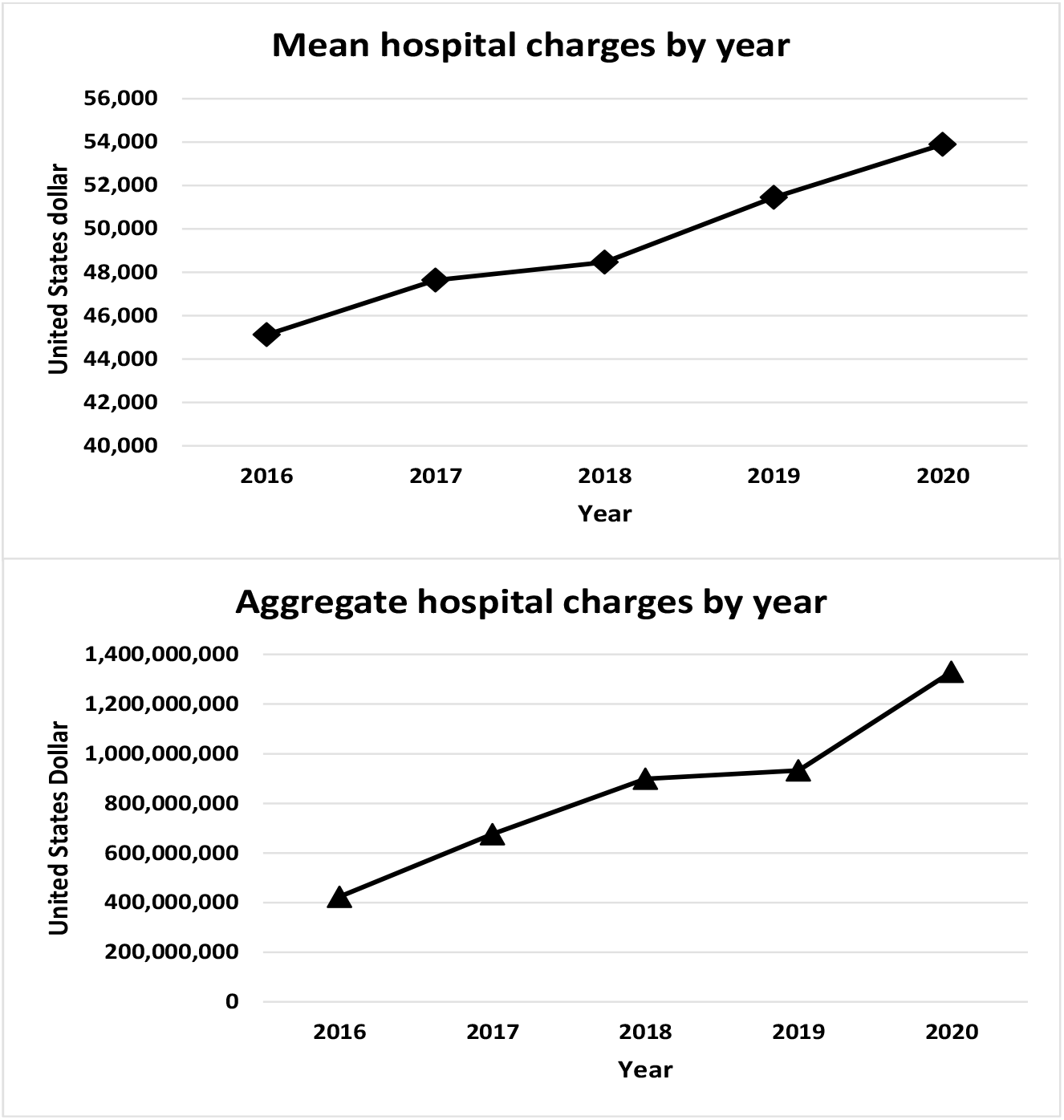
Trend in mean and aggregate hospital costs over five years.

### Patients discharge disposition

The majority of hospitalizations analyzed in the study resulted in routine home discharges, accounting for 54,368 cases, which represents approximately 80.6% of all admissions. Around 9.6% (6,476) of patients were discharged to skilled nursing facilities, intermediate care, or other types of specialized facilities. Additionally, 5.4% (3,643) of gender dysphoria individuals were discharged to home health care services. A smaller proportion of patients, about 1.3% (877), were discharged to short-term facilities, while 2.6% (1,754) chose to leave against medical advice.

Among the subgroup of gender dysphoria patients receiving gender-affirming care, the majority, 91%, were discharged routinely to their homes. A smaller percentage, 6.2%, required continued care through home health services. Furthermore, 2.4% of these patients were discharged to skilled nursing or intermediate care facilities to receive the necessary ongoing support and assistance.

## Discussion

The study analyzed 67,454 hospital discharges of individuals with gender dysphoria receiving gender-affirming care in the United States. There was a slight majority of individuals assigned female at birth (53.6%) compared to those assigned male at birth (46.4%). The mean age of the study population was 33 years, indicating that the majority of patients seeking gender-affirming care were relatively young adults [15]. Most hospitalizations (51.6%) were concentrated in the age group of 16 to 35 years, which aligns with previous literature reporting that many individuals undergo gender exploration and identity development during young adulthood [16,17]. However, the study also noted that 9.5% of patients were 60 years or older, which suggests that gender-affirming care is sought by a diverse age range of individuals. Access to healthcare was generally well-supported in this population with the majority of patients (94.3%) having insurance coverage. Additionally, the fact that hospitalizations were evenly distributed across all quartiles of median annual income suggests that gender-affirming care is being accessed across different socioeconomic groups.

The study highlights the types of services sought by individuals with gender dysphoria during hospitalizations. Mental health services were the most commonly provided services, accounting for approximately 55.2% of all admissions, which is consistent with prior research highlighting the high prevalence of depression and the importance of mental health support during gender transition [18,19].

Cross-sex hormonal therapy was another prevalent service, received by around 48.8% of individuals. This treatment aligns with established clinical guidelines [20], and its relatively high utilization suggests that hormone therapy is a significant component of gender-affirming care. Gender-affirming surgeries were performed in 7.1% of hospitalizations, indicating that a considerable number of individuals undergo surgical interventions to align their physical characteristics with their gender identity. The index study further reveals that surgeries were most frequently performed on individuals aged 26 to 35, followed by those in the age range of 16 to 25. Additionally, individuals aged 60 and above also underwent gender-affirming surgeries, highlighting that gender transition is not limited to younger age groups.

The study investigated various outcome measures related to hospitalization, including hospital length of stay, mortality, and total hospital charges. The average length of hospital stay for individuals with gender dysphoria was 6 days. Patients in the age range of 5 to 15 years had longer hospital stays compared to other age groups. This finding might be attributed to the need for additional medical and psychological support during gender exploration and treatment decisions for younger patients [21,22].

The study reported a total of 355 mortalities among individuals with gender dysphoria during the study period. Mortality rates were highest among individuals over 60 years old, which could be influenced by the burden of comorbidities and overall higher mortality rates among older populations. No mortality was recorded among individuals who received gender-affirming surgery, suggesting that this aspect of gender-affirming care may not significantly increase mortality risk in the short term. However, current evidence suggests an increased long-term risk of suicidal ideation and mortality after completion of sex reassignment [23]. The study’s reported mortality rates among different racial groups are essential for understanding disparities in healthcare outcomes for gender dysphoria and may contribute to ongoing efforts to reduce health disparities based on race and ethnicity.

The total hospital costs for all admissions of individuals with gender dysphoria during the five-year study period amounted to 3.33 billion US$, with an average cost of $49,830 per hospitalization. More money was allocated to other transition-related services and general care compared to gender-affirming procedures. This finding could potentially be influenced by the higher frequency of mental health services and cross-sex hormonal therapy, which were more commonly provided across all age groups. The increase in the mean and aggregate costs of hospitalization over time, suggests that gender-affirming care is becoming more widely accepted and accessible, leading to increased healthcare utilization and costs. These findings are consistent with the growing awareness and acceptance of gender diversity, leading to an increase in the demand for gender-affirming care.

Overall, the findings of this study align with existing literature on gender dysphoria and gender-affirming care. The predominance of young adults seeking gender-affirming care is consistent with previous research, as is the high utilization of mental health services and hormone therapy during gender transition.

The index study has some limitations that should be acknowledged. Firstly, the study’s data are limited to hospitalizations, which may not capture all instances of gender-affirming care sought by gender dysphoria individuals. Outpatient care, which can also be an integral part of gender-affirming treatment, is not included in the present analysis. Secondly, the study does not provide information on specific diagnoses, long-term outcomes, and follow-up care, which are essential for assessing the overall effectiveness and safety of gender-affirming interventions.

## Conclusion

In current study provides useful insights into hospitalization patterns and outcomes for individuals with gender dysphoria receiving gender-affirming care in the United States. Mental health services and cross-sex hormonal therapy were the most commonly provided during hospitalizations, emphasizing their crucial role in gender transition. Gender-affirming surgeries were performed on individuals across diverse age groups, including those aged 60 and above, with no recorded mortality, suggesting a relatively low mortality risk associated with this care aspect. Nonetheless, vigilance is necessary to address healthcare disparities, especially for individuals over 60 years old. The substantial total hospital costs underscore the significance of gender-affirming care, particularly in mental health services and hormone therapy. Our findings highlight the need for continued research and comprehensive support to meet the evolving healthcare needs of this important patient population, advancing gender-affirming care provision and understanding.

## Ethical considerations

This study utilized de-identified data from the NIS database, which is compliant with the Health Insurance Portability and Accountability Act (HIPAA) regulations. The NIS excludes all direct patient and hospital-level identifiers to ensure confidentiality. Further confidentiality measures include the removal of state identifiers and data elements that vary across states, such as hospital identifiers, secondary payer information, and state-specific coding. The utilization of the NIS in this study did not require review by an institutional review board. For this type of study, formal informed consent is not required.

## Data availability statement

All NIS datasets are publicly available through the HCUP central distributor upon request at https://hcup-us.ahrq.gov/tech_assist/centdist.jsp

## Funding

This study did not receive any funding

### Informed consent

The absence of patient and hospital-level identifiers in the NIS precludes obtaining informed consent. Nevertheless, this study diligently adheres to the standards mandated by the Agency for Healthcare Research and Quality (AHRQ), which effectively manages and regulates the use of the Nationwide Inpatient Sample.

### Studies involving human participants, their data, or biological material

This article does not contain any studies with human participants or animals performed by any of the authors.”

Informed consent

## Author contributions

F. Uwumiro: Conceptualization, Data curation, Formal Analysis, Writing (Original Draft & Review), Approval.

M. Bojerenu: Project supervision, Formal Analysis, Writing (Original Draft & Review), Approval.

C. Obijuru: Formal Analysis, Writing (Original Draft & Review), Approval.

O. Okunlola: Conceptualization, Writing (Original Draft & Review), Approval.

V. Nwike: Project supervision, Resources, Conceptualization, Writing (Original Draft & Review), Approval.

S. Abdul: Data Interpretation, Validation, Writing (Original Draft & Review), Approval.

H. Yayehyirad: Data Interpretation, Writing (Original Draft & Review), Approval.

S. Udegbe: Writing (Original Draft & Review), Approval.

O. Abesin: Data Interpretation, Writing (Original Draft & Review), Approval.

G. Orjih: Writing (Original Draft & Review), Visualization, Approval.

