## Appendix for "Hospitalization Patterns, Outcomes, and Resource Utilization among Individuals with Gender Dysphoria: A Nationwide Study"

**ICD-10 DIAGNOSIS CODES FOR GENDER DYSPHORIA USED IN THE STUDY**

| **ICD-10-CM** | **Code Description** |
| --- | --- |
| F64.0 | Transsexualism |
| F64.1 | Dual role transvestism |
| F64.2 | Gender identity disorder of childhood |
| F64.8 | Other gender identity disorders |
| F64.9 | Gender identity disorder, unspecified |
| Z87.890 | Personal history of sex reassignment |

**ICD-10 Procedure Codes**

**Male-to-Female Surgery**

| **ICD-10-PCS**  **procedure codes:** | **Code Description** |
| --- | --- |
| 0VTC0ZZ | Resection of Bilateral Testes, Open Approach |
| 0H0T0ZZ | Alteration of Right Breast, Open Approach |
| 0H0T3ZZ | Alteration of Right Breast, Percutaneous Approach |
| 0H0TXZZ | Alteration of Right Breast, External Approach |
| 0H0U0ZZ | Alteration of Left Breast, Open Approach |
| 0H0U3ZZ | Alteration of Left Breast, Percutaneous Approach |
| 0H0UXZZ | Alteration of Left Breast, External Approach |
| 0H0V07Z | Alteration of Bilateral Breast with Autologous Tissue Substitute, Open Approach |
| 0H0V0JZ | Alteration of Bilateral Breast with Synthetic Substitute, Open Approach |
| 0H0V0KZ | Alteration of Bilateral Breast with Nonautologous Tissue Substitute, Open Approach |
| 0H0V0ZZ | Alteration of Bilateral Breast, Open Approach |
| 0H0V37Z | Alteration of Bilateral Breast with Autologous Tissue Substitute, Percutaneous Approach |
| 0H0V3JZ | Alteration of Bilateral Breast with Synthetic Substitute, Percutaneous Approach |
| 0H0V3KZ | Alteration of Bilateral Breast with Nonautologous Tissue Substitute, Percutaneous Approach |
| 0H0V3ZZ | Alteration of Bilateral Breast, Percutaneous Approach |
| 0H0VXZZ | Alteration of Bilateral Breast, External Approach |
| 0HMTXZZ | Reattachment of Right Breast, External Approach |
| 0HMUXZZ | Reattachment of Left Breast, External Approach |
| 0HMVXZZ | Reattachment of Bilateral Breast, External Approach |
| 0HMWXZZ | Reattachment of Right Nipple, External Approach |
| 0HMXXZZ | Reattachment of Left Nipple, External Approach |
| 0HNT0ZZ | Release Right Breast, Open Approach |
| 0HNT3ZZ | Release Right Breast, Percutaneous Approach |
| 0HNT7ZZ | Release Right Breast, Via Natural or Artificial Opening |
| 0HNT8ZZ | Release Right Breast, Via Natural or Artificial Opening Endoscopic |
| 0HNTXZZ | Release Right Breast, External Approach |
| 0HNU0ZZ | Release Left Breast, Open Approach |
| 0HNU3ZZ | Release Left Breast, Percutaneous Approach |
| 0HNU7ZZ | Release Left Breast, Via Natural or Artificial Opening |
| 0HNU8ZZ | Release Left Breast, Via Natural or Artificial Opening Endoscopic |
| 0HNUXZZ | Release Left Breast, External Approach |
| 0HNV0ZZ | Release Bilateral Breast, Open Approach |
| 0HNV3ZZ | Release Bilateral Breast, Percutaneous Approach |
| 0HNV7ZZ | Release Bilateral Breast, Via Natural or Artificial Opening |
| 0HNV8ZZ | Release Bilateral Breast, Via Natural or Artificial Opening Endoscopic |
| 0HNVXZZ | Release Bilateral Breast, External Approach |
| 0HNW0ZZ | Release Right Nipple, Open Approach |
| 0HNW3ZZ | Release Right Nipple, Percutaneous Approach |
| 0HNW7ZZ | Release Right Nipple, Via Natural or Artificial Opening |
| 0HNW8ZZ | Release Right Nipple, Via Natural or Artificial Opening Endoscopic |
| 0HNWXZZ | Release Right Nipple, External Approach |
| 0HNX0ZZ | Release Left Nipple, Open Approach |
| 0HNX3ZZ | Release Left Nipple, Percutaneous Approach |
| 0HNX7ZZ | Release Left Nipple, Via Natural or Artificial Opening |
| 0HNX8ZZ | Release Left Nipple, Via Natural or Artificial Opening Endoscopic |
| 0HNXXZZ | Release Left Nipple, External Approach |
| 0HQT0ZZ | Repair Right Breast, Open Approach |
| 0HQT3ZZ | Repair Right Breast, Percutaneous Approach |
| 0HQT7ZZ | Repair Right Breast, Via Natural or Artificial Opening |
| 0HQT8ZZ | Repair Right Breast, Via Natural or Artificial Opening Endoscopic |
| 0HQTXZZ | Repair Right Breast, External Approach |
| 0HQU0ZZ | Repair Left Breast, Open Approach |
| 0HQU3ZZ | Repair Left Breast, Percutaneous Approach |
| 0HQU3ZZ | Repair Left Breast, Via Natural or Artificial Opening |
| 0HQU8ZZ | Repair Left Breast, Via Natural or Artificial Opening Endoscopic |
| 0HQUXZZ | Repair Left Breast, External Approach |
| 0HQV0ZZ | Repair Bilateral Breast, Open Approach |
| 0HQV3ZZ | Repair Bilateral Breast, Percutaneous Approach |
| 0HQV7ZZ | Repair Bilateral Breast, Via Natural or Artificial Opening |
| 0HQV8ZZ | Repair Bilateral Breast, Via Natural or Artificial Opening Endoscopic |
| 0HQVXZZ | Repair Bilateral Breast, External Approach |
| 0HQW0ZZ | Repair Right Nipple, Open Approach |
| 0HQW3ZZ | Repair Right Nipple, Percutaneous Approach |
| 0HQW7ZZ | Repair Right Nipple, Via Natural or Artificial Opening |
| 0HQW8ZZ | Repair Right Nipple, Via Natural or Artificial Opening Endoscopic |
| 0HQWXZZ | Repair Right Nipple, External Approach |
| 0HQX0ZZ | Repair Left Nipple, Open Approach |
| 0HQX3ZZ | Repair Left Nipple, Percutaneous Approach |
| 0HQX7ZZ | Repair Left Nipple, Via Natural or Artificial Opening |
| 0HQX8ZZ | Repair Left Nipple, Via Natural or Artificial Opening Endoscopic |
| 0HQXXZZ | Repair Left Nipple, External Approach |
| 0HQY0ZZ | Repair Supernumerary Breast, Open Approach |
| 0HQY3ZZ | Repair Supernumerary Breast, Percutaneous Approach |
| 0HQY7ZZ | Repair Supernumerary Breast, Via Natural or Artificial Opening |
| 0HQY8ZZ | Repair Supernumerary Breast, Via Natural or Artificial Opening Endoscopic |
| 0HQYXZZ | Repair Supernumerary Breast, External Approach |
| 0HRT07Z | Replacement of Right Breast with Autologous Tissue Substitute, Open Approach |
| 0HRT0KZ | Replacement of Right Breast with Nonautologous Tissue Substitute, Open Approach |
| 0HRT37Z | Replacement of Right Breast with Autologous Tissue Substitute, Percutaneous  Approach |
| 0HRT3KZ | Replacement of Right Breast with Nonautologous Tissue Substitute, Percutaneous  Approach |
| 0HRTXJZ | Replacement of Right Breast with Synthetic Substitute, External Approach |
| 0HRU07Z | Replacement of Left Breast with Autologous Tissue Substitute, Open Approach |
| 0HRU0KZ | Replacement of Left Breast with Nonautologous Tissue Substitute, Open Approach |
| 0HRU37Z | Replacement of Left Breast with Autologous Tissue Substitute, Percutaneous Approach |
| 0HRU3KZ | Replacement of Left Breast with Nonautologous Tissue Substitute, Percutaneous Approach |
| 0HRUXJZ | Replacement of Left Breast with Synthetic Substitute, External Approach |
| 0HRV07Z | Replacement of Bilateral Breast with Autologous Tissue Substitute, Open Approach |
| 0HRV0KZ | Replacement of Bilateral Breast with Nonautologous Tissue Substitute, Open Approach |
| 0HRV37Z | Replacement of Bilateral Breast with Autologous Tissue Substitute, Percutaneous Approach |
| 0HRV3KZ | Replacement of Bilateral Breast with Nonautologous Tissue Substitute, Percutaneous Approach |
| 0HRVXJZ | Replacement of Bilateral Breast with Synthetic Substitute, External Approach |
| 0HRW07Z | Replacement of Right Nipple with Autologous Tissue Substitute, Open Approach |
| 0HRW0JZ | Replacement of Right Nipple with Synthetic Substitute, Open Approach |
| 0HRW0KZ | Replacement of Right Nipple with Nonautologous Tissue Substitute, Open Approach |
| 0HRW37Z | Replacement of Right Nipple with Autologous Tissue Substitute, Percutaneous Approach |
| 0HRW3JZ | Replacement of Right Nipple with Synthetic Substitute, Percutaneous Approach |
| 0HRW3KZ | Replacement of Right Nipple with Nonautologous Tissue Substitute, Percutaneous Approach |
| 0HRWX7Z | Replacement of Right Nipple with Autologous Tissue Substitute, External Approach |
| 0HRWXJZ | Replacement of Right Nipple with Synthetic Substitute, External Approach |
| 0HRWXKZ | Replacement of Right Nipple with Nonautologous Tissue Substitute, External Approach |
| 0HRX07Z | Replacement of Left Nipple with Autologous Tissue Substitute, Open Approach |
| 0HRX0JZ | Replacement of Left Nipple with Synthetic Substitute, Open Approach |
| 0HRX0KZ | Replacement of Left Nipple with Nonautologous Tissue Substitute, Open Approach |
| 0HRX37Z | Replacement of Left Nipple with Autologous Tissue Substitute, Percutaneous Approach |
| 0HRX3JZ | Replacement of Left Nipple with Synthetic Substitute, Percutaneous Approach |
| 0HRX3KZ | Replacement of Left Nipple with Nonautologous Tissue Substitute, Percutaneous  Approach |
| 0HRXX7Z | Replacement of Left Nipple with Autologous Tissue Substitute, External Approach |
| 0HRXXJZ | Replacement of Left Nipple with Synthetic Substitute, External Approach |
| 0HRXXKZ | Replacement of Left Nipple with Nonautologous Tissue Substitute, External Approach |
| 0HUT07Z | Supplement Right Breast with Autologous Tissue Substitute, Open Approach |
| 0HUT0JZ | Supplement Right Breast with Synthetic Substitute, Open Approach |
| 0HUT0KZ | Supplement Right Breast with Nonautologous Tissue Substitute, Open Approach |
| 0HUT37Z | Supplement Right Breast with Autologous Tissue Substitute, Percutaneous Approach |
| 0HUT3JZ | Supplement Right Breast with Synthetic Substitute, Percutaneous Approach |
| 0HUT3KZ | Supplement Right Breast with Nonautologous Tissue Substitute, Percutaneous Approach |
| 0HUT77Z | Supplement Right Breast with Autologous Tissue Substitute, Via Natural or Artificial  Opening |
| 0HUT7JZ | Supplement Right Breast with Synthetic Substitute, Via Natural or Artificial Opening |
| 0HUT7KZ | Supplement Right Breast with Nonautologous Tissue Substitute, Via Natural or Artificial Opening |
| 0HUT87Z | Supplement Right Breast with Autologous Tissue Substitute, Via Natural or Artificial Opening Endoscopic |
| 0HUT8JZ | Supplement Right Breast with Synthetic Substitute, Via Natural or Artificial Opening  Endoscopic |
| 0HUT8KZ | Supplement Right Breast with Nonautologous Tissue Substitute, Via Natural or Artificial  Opening Endoscopic |
| 0HUTX7Z | Supplement Right Breast with Autologous Tissue Substitute, External Approach |
| 0HUTXJZ | Supplement Right Breast with Synthetic Substitute, External Approach |
| 0HUTXKZ | Supplement Right Breast with Nonautologous Tissue Substitute, External Approach |
| 0HUU07Z | Supplement Left Breast with Autologous Tissue Substitute, Open Approach |
| 0HUU0JZ | Supplement Left Breast with Synthetic Substitute, Open Approach |
| 0HUU0KZ | Supplement Left Breast with Nonautologous Tissue Substitute, Open Approach |
| 0HUU37Z | Supplement Left Breast with Autologous Tissue Substitute, Percutaneous Approach |
| 0HUU3JZ | Supplement Left Breast with Synthetic Substitute, Percutaneous Approach |
| 0HUU3KZ | Supplement Left Breast with Nonautologous Tissue Substitute, Percutaneous Approach |
| 0HUU77Z | Supplement Left Breast with Autologous Tissue Substitute, Via Natural or Artificial Opening |
| 0HUU7JZ | Supplement Left Breast with Synthetic Substitute, Via Natural or Artificial Opening |
| 0HUU7KZ | Supplement Left Breast with Nonautologous Tissue Substitute, Via Natural or Artificial  Opening |
| 0HUU87Z | Supplement Left Breast with Autologous Tissue Substitute, Via Natural or Artificial Opening Endoscopic |
| 0HUU8JZ | Supplement Left Breast with Synthetic Substitute, Via Natural or Artificial Opening Endoscopic |
| 0HUU8KZ | Supplement Left Breast with Nonautologous Tissue Substitute, Via Natural or Artificial Opening Endoscopic |
| 0HUUX7Z | Supplement Left Breast with Autologous Tissue Substitute, External Approach |
| 0HUUXJZ | Supplement Left Breast with Synthetic Substitute, External Approach |
| 0HUUXKZ | Supplement Left Breast with Nonautologous Tissue Substitute, External Approach |
| 0HUV07Z | Supplement Bilateral Breast with Autologous Tissue Substitute, Open Approach |
| 0HUV0JZ | Supplement Bilateral Breast with Synthetic Substitute, Open Approach |
| 0HUV0KZ | Supplement Bilateral Breast with Nonautologous Tissue Substitute, Open Approach |
| 0HUV37Z | Supplement Bilateral Breast with Autologous Tissue Substitute, Percutaneous  Approach |
| 0HUV3JZ | Supplement Bilateral Breast with Synthetic Substitute, Percutaneous Approach |
| 0HUV3KZ | Supplement Bilateral Breast with Nonautologous Tissue Substitute, Percutaneous  Approach |
| 0HUV77Z | Supplement Bilateral Breast with Autologous Tissue Substitute, Via Natural or Artificial  Opening |
| 0HUV7JZ | Supplement Bilateral Breast with Synthetic Substitute, Via Natural or Artificial Opening |
| 0HUV7KZ | Supplement Bilateral Breast with Nonautologous Tissue Substitute, Via Natural or Artificial Opening |
| 0HUV87Z | Supplement Bilateral Breast with Autologous Tissue Substitute, Via Natural or Artificial  Opening Endoscopic |
| 0HUV8JZ | Supplement Bilateral Breast with Synthetic Substitute, Via Natural or Artificial Opening  Endoscopic |
| 0HUV8KZ | Supplement Bilateral Breast with Nonautologous Tissue Substitute, Via Natural or  Artificial Opening Endoscopic |
| 0HUVX7Z | Supplement Bilateral Breast with Autologous Tissue Substitute, External Approach |
| 0HUVXJZ | Supplement Bilateral Breast with Synthetic Substitute, External Approach |
| 0HUVXKZ | Supplement Bilateral Breast with Nonautologous Tissue Substitute, External Approach |
| 0HUW07Z | Supplement Right Nipple with Autologous Tissue Substitute, Open Approach |
| 0HUW0JZ | Supplement Right Nipple with Synthetic Substitute, Open Approach |
| 0HUW0KZ | Supplement Right Nipple with Nonautologous Tissue Substitute, Open Approach |
| 0HUW37Z | Supplement Right Nipple with Autologous Tissue Substitute, Percutaneous Approach |
| 0HUW3JZ | Supplement Right Nipple with Synthetic Substitute, Percutaneous Approach |
| 0HUW3KZ | Supplement Right Nipple with Nonautologous Tissue Substitute, Percutaneous  Approach |
| 0HUW77Z | Supplement Right Nipple with Autologous Tissue Substitute, Via Natural or Artificial  Opening |
| 0HUW7JZ | Supplement Right Nipple with Synthetic Substitute, Via Natural or Artificial Opening |
| 0HUW7KZ | Supplement Right Nipple with Nonautologous Tissue Substitute, Via Natural or Artificial Opening |
| 0HUW87Z | Supplement Right Nipple with Autologous Tissue Substitute, Via Natural or Artificial  Opening Endoscopic |
| 0HUW8JZ | Supplement Right Nipple with Synthetic Substitute, Via Natural or Artificial Opening  Endoscopic |
| 0HUW8KZ | Supplement Right Nipple with Nonautologous Tissue Substitute, Via Natural or Artificial  Opening Endoscopic |
| 0HUWX7Z | Supplement Right Nipple with Autologous Tissue Substitute, External Approach |
| 0HUWXJZ | Supplement Right Nipple with Synthetic Substitute, External Approach |
| 0HUWXKZ | Supplement Right Nipple with Nonautologous Tissue Substitute, External Approach |
| 0HUX07Z | Supplement Left Nipple with Autologous Tissue Substitute, Open Approach |
| 0HUX0JZ | Supplement Left Nipple with Synthetic Substitute, Open Approach |
| 0HUX0KZ | Supplement Left Nipple with Nonautologous Tissue Substitute, Open Approach |
| 0HUX37Z | Supplement Left Nipple with Autologous Tissue Substitute, Percutaneous Approach |
| 0HUX3JZ | Supplement Left Nipple with Synthetic Substitute, Percutaneous Approach |
| 0HUX3KZ | Supplement Left Nipple with Nonautologous Tissue Substitute, Percutaneous Approach |
| 0HUX77Z | Supplement Left Nipple with Autologous Tissue Substitute, Via Natural or Artificial Opening |
| 0HUX7JZ | Supplement Left Nipple with Synthetic Substitute, Via Natural or Artificial Opening |
| 0HUX7KZ | Supplement Left Nipple with Nonautologous Tissue Substitute, Via Natural or Artificial  Opening |
| 0HUX87Z | Supplement Left Nipple with Autologous Tissue Substitute, Via Natural or Artificial  Opening Endoscopic |
| 0HUX8JZ | Supplement Left Nipple with Synthetic Substitute, Via Natural or Artificial Opening Endoscopic |
| 0HUX8KZ | Supplement Left Nipple with Nonautologous Tissue Substitute, Via Natural or Artificial Opening Endoscopic |
| 0HUXX7Z | Supplement Left Nipple with Autologous Tissue Substitute, External Approach |
| 0HUXXJZ | Supplement Left Nipple with Synthetic Substitute, External Approach |
| 0HUXXKZ | Supplement Left Nipple with Nonautologous Tissue Substitute, External Approach |
| 0U5J0ZZ | Destruction of Clitoris, Open Approach |
| 0U5JXZZ | Destruction of Clitoris, External Approach |
| 0U9J00Z | Drainage of Clitoris with Drainage Device, Open Approach |
| 0U9J0ZZ | Drainage of Clitoris, Open Approach |
| 0U9JX0Z | Drainage of Clitoris with Drainage Device, External Approach |
| 0U9JXZZ | Drainage of Clitoris, External Approach |
| 0UBJ0ZX | Excision of Clitoris, Open Approach, Diagnostic |
| 0UBJ0ZZ | Excision of Clitoris, Open Approach |
| 0UBJXZX | Excision of Clitoris, External Approach, Diagnostic |
| 0UBJXZZ | Excision of Clitoris, External Approach |
| 0UCJ0ZZ | Extirpation of Matter from Clitoris, Open Approach |
| 0UCJXZZ | Extirpation of Matter from Clitoris, External Approach |
| 0UMJXZZ | Reattachment of Clitoris, External Approach |
| 0UNJ0ZZ | Release Clitoris, Open Approach |
| 0UNJXZZ | Release Clitoris, External Approach |
| 0UQG0ZZ | Repair Vagina, Open Approach |
| 0UQJ0ZZ | Repair Clitoris, Open Approach |
| 0UQJXZZ | Repair Clitoris, External Approach |
| 0UTJ0ZZ | Resection of Clitoris, Open Approach |
| 0UTJXZZ | Resection of Clitoris, External Approach |
| 0UUG07Z | Supplement Vagina with Autologous Tissue Substitute, Open Approach |
| 0UUG0JZ | Supplement Vagina with Synthetic Substitute, Open Approach |
| 0UUG0KZ | Supplement Vagina with Nonautologous Tissue Substitute, Open Approach |
| 0UUG47Z | Supplement Vagina with Autologous Tissue Substitute, Percutaneous Endoscopic Approach |
| 0UUG4JZ | Supplement Vagina with Synthetic Substitute, Percutaneous Endoscopic Approach |
| 0UUG4KZ | Supplement Vagina with Nonautologous Tissue Substitute, Percutaneous Endoscopic Approach |
| 0UUG77Z | Supplement Vagina with Autologous Tissue Substitute, Via Natural or Artificial Opening |
| 0UUG7JZ | Supplement Vagina with Synthetic Substitute, Via Natural or Artificial Opening |
| 0UUG7KZ | Supplement Vagina with Nonautologous Tissue Substitute, Via Natural or Artificial |
|  | Opening |
| 0UUG87Z | Supplement Vagina with Autologous Tissue Substitute, Via Natural or Artificial Opening Endoscopic |
| 0UUG8JZ | Supplement Vagina with Synthetic Substitute, Via Natural or Artificial Opening  Endoscopic |
| 0UUG8KZ | Supplement Vagina with Nonautologous Tissue Substitute, Via Natural or Artificial  Opening Endoscopic |
| 0UUGX7Z | Supplement Vagina with Autologous Tissue Substitute, External Approach |
| 0UUGXJZ | Supplement Vagina with Synthetic Substitute, External Approach |
| 0UUGXKZ | Supplement Vagina with Nonautologous Tissue Substitute, External Approach |
| 0UUJ07Z | Supplement Clitoris with Autologous Tissue Substitute, Open Approach |
| 0UUJ0JZ | Supplement Clitoris with Synthetic Substitute, Open Approach |
| 0UUJ0KZ | Supplement Clitoris with Nonautologous Tissue Substitute, Open Approach |
| 0UUJX7Z | Supplement Clitoris with Autologous Tissue Substitute, External Approach |
| 0UUJXJZ | Supplement Clitoris with Synthetic Substitute, External Approach |
| 0UUJXKZ | Supplement Clitoris with Nonautologous Tissue Substitute, External Approach |
| 0VT90ZZ | Resection of Right Testis, Open Approach |
| 0VT94ZZ | Resection of Right Testis, Percutaneous Endoscopic Approach |
| 0VTB0ZZ | Resection of Left Testis, Open Approach |
| 0VTB4ZZ | Resection of Left Testis, Percutaneous Endoscopic Approach |
| 0VTC4ZZ | Resection of Bilateral Testes, Percutaneous Endoscopic Approach |
| 0VTS0ZZ | Resection of Penis, Open Approach |
| 0VTS4ZZ | Resection of Penis, Percutaneous Endoscopic Approach |
| 0VTSXZZ | Resection of Penis, External Approach |
| 0W4M070 | Creation of Vagina in Male Perineum with Autologous Tissue Substitute, Open  Approach |
| 0W4M0J0 | Creation of Vagina in Male Perineum with Synthetic Substitute, Open Approach |
| 0W4M0K0 | Creation of Vagina in Male Perineum with Nonautologous Tissue Substitute, Open Approach |
| 0W4M0Z0 | Creation of Vagina in Male Perineum, Open Approach |

**Male-to-Female Facial Feminization**

|  | **ICD-10-PCS**  **procedure codes:** | | **Code Description** |
| --- | --- | --- | --- |
|  | 08SN0ZZ | | Reposition Right Upper Eyelid, Open Approach |
|  | 08SN3ZZ | | Reposition Right Upper Eyelid, Percutaneous Approach |
|  | 08SNXZZ | | Reposition Right Upper Eyelid, External Approach |
|  | 08SP0ZZ | | Reposition Left Upper Eyelid, Open Approach |
|  | 08SP3ZZ | | Reposition Left Upper Eyelid, Percutaneous Approach |
|  | 08SPXZZ | | Reposition Left Upper Eyelid, External Approach |
|  | 08SQ0ZZ | | Reposition Right Lower Eyelid, Open Approach |
|  | 08SQ3ZZ | | Reposition Right Lower Eyelid, Percutaneous Approach |
|  | 08SQXZZ | | Reposition Right Lower Eyelid, External Approach |
|  | 08SR0ZZ | | Reposition Left Lower Eyelid, Open Approach |
|  | 08SR3ZZ | | Reposition Left Lower Eyelid, Percutaneous Approach |
|  | 08SRXZZ | | Reposition Left Lower Eyelid, External Approach |
|  | 0HD0XZZ | | Extraction of Scalp Skin, External Approach |
|  | 0HD1XZZ | | Extraction of Face Skin, External Approach |
|  | 0HD4XZZ | | Extraction of Neck Skin, External Approach |
|  | 0HD5XZZ | | Extraction of Chest Skin, External Approach |
|  | 0HD6XZZ | | Extraction of Back Skin, External Approach |
|  | 0HD7XZZ | | Extraction of Abdomen Skin, External Approach |
|  | 0HD8XZZ | | Extraction of Buttock Skin, External Approach |
|  | 0HDAXZZ | | Extraction of Genitalia Skin, External Approach |
|  | 0HDBXZZ | | Extraction of Right Upper Arm Skin, External Approach |
|  | 0HDCXZZ | | Extraction of Left Upper Arm Skin, External Approach |
|  | 0HDDXZZ | | Extraction of Right Lower Arm Skin, External Approach |
|  | 0HDEXZZ | | Extraction of Left Lower Arm Skin, External Approach |
|  | 0HDFXZZ | | Extraction of Right Hand Skin, External Approach |
|  | 0HDGXZZ | | Extraction of Left Hand Skin, External Approach |
|  | 0HDHXZZ | | Extraction of Right Upper Leg Skin, External Approach |
|  | 0HDJXZZ | | Extraction of Left Upper Leg Skin, External Approach |
|  | 0HDKXZZ | | Extraction of Right Lower Leg Skin, External Approach |
|  | 0HDLXZZ | | Extraction of Left Lower Leg Skin, External Approach |
|  | 0HDMXZZ | | Extraction of Right Foot Skin, External Approach |
|  | 0HDNXZZ | | Extraction of Left Foot Skin, External Approach |
|  | 0JBL0ZZ | | Excision of Right Upper Leg Subcutaneous Tissue and Fascia, Open Approach |
|  | 0JBL3ZZ | | Excision of Right Upper Leg Subcutaneous Tissue and Fascia, Percutaneous Approach |
|  | 0JBM0ZZ | | Excision of Left Upper Leg Subcutaneous Tissue and Fascia, Open Approach |
|  | 0JBM3ZZ | | Excision of Left Upper Leg Subcutaneous Tissue and Fascia, Percutaneous Approach |
|  | 0KS10ZZ | | Reposition Facial Muscle, Open Approach |
|  | 0KS14ZZ | | Reposition Facial Muscle, Percutaneous Endoscopic Approach |
|  | 0NNC0ZZ | | Release Right Sphenoid Bone, Open Approach |
|  | 0NNC3ZZ | | Release Right Sphenoid Bone, Percutaneous Approach |
|  | 0NNC4ZZ | | Release Right Sphenoid Bone, Percutaneous Endoscopic Approach |
|  | 0NND0ZZ | | Release Left Sphenoid Bone, Open Approach |
|  | 0NND3ZZ | | Release Left Sphenoid Bone, Percutaneous Approach |
|  | 0NND4ZZ | | Release Left Sphenoid Bone, Percutaneous Endoscopic Approach |
|  | 0NNF0ZZ | | Release Right Ethmoid Bone, Open Approach |
|  | 0NNF3ZZ | | Release Right Ethmoid Bone, Percutaneous Approach |
|  | 0NNF4ZZ | | Release Right Ethmoid Bone, Percutaneous Endoscopic Approach |
|  | 0NNG0ZZ | | Release Left Ethmoid Bone, Open Approach |
|  | 0NNG3ZZ | | Release Left Ethmoid Bone, Percutaneous Approach |
|  | 0NNG4ZZ | | Release Left Ethmoid Bone, Percutaneous Endoscopic Approach |
|  | 0NNH0ZZ | | Release Right Lacrimal Bone, Open Approach |
|  | 0NNH3ZZ | | Release Right Lacrimal Bone, Percutaneous Approach |
|  | 0NNH4ZZ | | Release Right Lacrimal Bone, Percutaneous Endoscopic Approach |
|  | 0NNJ0ZZ | | Release Left Lacrimal Bone, Open Approach |
|  | 0NNJ3ZZ | | Release Left Lacrimal Bone, Percutaneous Approach |
|  | 0NNJ4ZZ | | Release Left Lacrimal Bone, Percutaneous Endoscopic Approach |
|  | 0NNK0ZZ | | Release Right Palatine Bone, Open Approach |
|  | 0NNK3ZZ | | Release Right Palatine Bone, Percutaneous Approach |
|  | 0NNK4ZZ | | Release Right Palatine Bone, Percutaneous Endoscopic Approach |
|  | 0NNL0ZZ | | Release Left Palatine Bone, Open Approach |
|  | 0NNL3ZZ | | Release Left Palatine Bone, Percutaneous Approach |
|  | 0NNL4ZZ | | Release Left Palatine Bone, Percutaneous Endoscopic Approach |
|  | 0NNM0ZZ | | Release Right Zygomatic Bone, Open Approach |
|  | 0NNM3ZZ | | Release Right Zygomatic Bone, Percutaneous Approach |
|  | 0NNM4ZZ | | Release Right Zygomatic Bone, Percutaneous Endoscopic Approach |
|  | 0NNN0ZZ | | Release Left Zygomatic Bone, Open Approach |
|  | 0NNN3ZZ | | Release Left Zygomatic Bone, Percutaneous Approach |
|  | 0NNN4ZZ | | Release Left Zygomatic Bone, Percutaneous Endoscopic Approach |
|  | 0NNP0ZZ | | Release Right Orbit, Open Approach |
|  | 0NNP3ZZ | | Release Right Orbit, Percutaneous Approach |
|  | 0NNP4ZZ | | Release Right Orbit, Percutaneous Endoscopic Approach |
|  | 0NNQ0ZZ | | Release Left Orbit, Open Approach |
|  | 0NNQ3ZZ | | Release Left Orbit, Percutaneous Approach |
|  | 0NNQ4ZZ | | Release Left Orbit, Percutaneous Endoscopic Approach |
|  | 0NNR0ZZ | | Release Right Maxilla, Open Approach |
|  | 0NNR3ZZ | | Release Right Maxilla, Percutaneous Approach |
|  | 0NNR4ZZ | | Release Right Maxilla, Percutaneous Endoscopic Approach |
|  | 0NNS0ZZ | | Release Left Maxilla, Open Approach |
|  | 0NNS3ZZ | | Release Left Maxilla, Percutaneous Approach |
|  | 0NNS4ZZ | | Release Left Maxilla, Percutaneous Endoscopic Approach |
|  | 0NNT0ZZ | | Release Right Mandible, Open Approach |
|  | 0NNT3ZZ | | Release Right Mandible, Percutaneous Approach |
|  | 0NNT4ZZ | | Release Right Mandible, Percutaneous Endoscopic Approach |
|  | 0NNV0ZZ | | Release Left Mandible, Open Approach |
|  | 0NNV3ZZ | | Release Left Mandible, Percutaneous Approach |
|  | 0NNV4ZZ | | Release Left Mandible, Percutaneous Endoscopic Approach |
|  | 0NQC0ZZ | | Repair Right Sphenoid Bone, Open Approach |
|  | 0NQC3ZZ | | Repair Right Sphenoid Bone, Percutaneous Approach |
|  | 0NQC4ZZ | | Repair Right Sphenoid Bone, Percutaneous Endoscopic Approach |
|  | 0NQCXZZ | | Repair Right Sphenoid Bone, External Approach |
|  | 0NQD0ZZ | | Repair Left Sphenoid Bone, Open Approach |
|  | 0NQD3ZZ | | Repair Left Sphenoid Bone, Percutaneous Approach |
|  | 0NQD4ZZ | | Repair Left Sphenoid Bone, Percutaneous Endoscopic Approach |
|  | 0NQDXZZ | | Repair Left Sphenoid Bone, External Approach |
|  | 0NQF0ZZ | | Repair Right Ethmoid Bone, Open Approach |
|  | 0NQF3ZZ | | Repair Right Ethmoid Bone, Percutaneous Approach |
|  | 0NQF4ZZ | | Repair Right Ethmoid Bone, Percutaneous Endoscopic Approach |
|  | 0NQFXZZ | | Repair Right Ethmoid Bone, External Approach |
|  | 0NQG0ZZ | | Repair Left Ethmoid Bone, Open Approach |
|  | 0NQG3ZZ | | Repair Left Ethmoid Bone, Percutaneous Approach |
|  | 0NQG4ZZ | | Repair Left Ethmoid Bone, Percutaneous Endoscopic Approach |
|  | 0NQGXZZ | | Repair Left Ethmoid Bone, External Approach |
|  | 0NQH0ZZ | | Repair Right Lacrimal Bone, Open Approach |
|  | 0NQH3ZZ | | Repair Right Lacrimal Bone, Percutaneous Approach |
|  | 0NQH4ZZ | | Repair Right Lacrimal Bone, Percutaneous Endoscopic Approach |
|  | 0NQHXZZ | | Repair Right Lacrimal Bone, External Approach |
|  | 0NQJ0ZZ | | Repair Left Lacrimal Bone, Open Approach |
|  | 0NQJ3ZZ | | Repair Left Lacrimal Bone, Percutaneous Approach |
|  | 0NQJ4ZZ | | Repair Left Lacrimal Bone, Percutaneous Endoscopic Approach |
|  | 0NQJXZZ | | Repair Left Lacrimal Bone, External Approach |
|  | 0NQK0ZZ | | Repair Right Palatine Bone, Open Approach |
|  | 0NQK3ZZ | | Repair Right Palatine Bone, Percutaneous Approach |
|  | 0NQK4ZZ | | Repair Right Palatine Bone, Percutaneous Endoscopic Approach |
|  | 0NQKXZZ | | Repair Right Palatine Bone, External Approach |
|  | 0NQL0ZZ | | Repair Left Palatine Bone, Open Approach |
|  | 0NQL3ZZ | | Repair Left Palatine Bone, Percutaneous Approach |
|  | 0NQL4ZZ | | Repair Left Palatine Bone, Percutaneous Endoscopic Approach |
|  | 0NQLXZZ | | Repair Left Palatine Bone, External Approach |
|  | 0NQM0ZZ | | Repair Right Zygomatic Bone, Open Approach |
|  | 0NQM3ZZ | | Repair Right Zygomatic Bone, Percutaneous Approach |
|  | 0NQM4ZZ | | Repair Right Zygomatic Bone, Percutaneous Endoscopic Approach |
|  | 0NQMXZZ | | Repair Right Zygomatic Bone, External Approach |
|  | 0NQN0ZZ | | Repair Left Zygomatic Bone, Open Approach |
|  | 0NQN3ZZ | | Repair Left Zygomatic Bone, Percutaneous Approach |
|  | 0NQN4ZZ | | Repair Left Zygomatic Bone, Percutaneous Endoscopic Approach |
|  | 0NQNXZZ | | Repair Left Zygomatic Bone, External Approach |
|  | 0NQX0ZZ | | Repair Hyoid Bone, Open Approach |
|  | 0NQX3ZZ | | Repair Hyoid Bone, Percutaneous Approach |
|  | 0NQX4ZZ | | Repair Hyoid Bone, Percutaneous Endoscopic Approach |
|  | 0NQXXZZ | | Repair Hyoid Bone, External Approach |
|  | 0NRC07Z | | Replacement of Right Sphenoid Bone with Autologous Tissue Substitute, Open Approach |
|  | 0NRC0JZ | | Replacement of Right Sphenoid Bone with Synthetic Substitute, Open Approach |
|  | 0NRC0KZ | | Replacement of Right Sphenoid Bone with Nonautologous Tissue Substitute, Open Approach |
|  | 0NRC37Z | | Replacement of Right Sphenoid Bone with Autologous Tissue Substitute, Percutaneous Approach |
|  | 0NRC3JZ | | Replacement of Right Sphenoid Bone with Synthetic Substitute, Percutaneous Approach |
|  | 0NRC3KZ | | Replacement of Right Sphenoid Bone with Nonautologous Tissue Substitute, Percutaneous Approach |
|  | 0NRC47Z | | Replacement of Right Sphenoid Bone with Autologous Tissue Substitute, Percutaneous Endoscopic Approach |
|  | 0NRC4JZ | | Replacement of Right Sphenoid Bone with Synthetic Substitute, Percutaneous  Endoscopic Approach |
|  | 0NRC4KZ | | Replacement of Right Sphenoid Bone with Nonautologous Tissue Substitute,  Percutaneous Endoscopic Approach |
|  | 0NRD07Z | | Replacement of Left Sphenoid Bone with Autologous Tissue Substitute, Open  Approach |
|  | 0NRD0JZ | | Replacement of Left Sphenoid Bone with Synthetic Substitute, Open Approach |
|  | 0NRD0KZ | | Replacement of Left Sphenoid Bone with Nonautologous Tissue Substitute, Open  Approach |
|  | 0NRD37Z | | Replacement of Left Sphenoid Bone with Autologous Tissue Substitute, Percutaneous  Approach |
|  | 0NRD3JZ | | Replacement of Left Sphenoid Bone with Synthetic Substitute, Percutaneous Approach |
|  | 0NRD3KZ | | Replacement of Left Sphenoid Bone with Nonautologous Tissue Substitute, Percutaneous Approach |
|  | 0NRD47Z | | Replacement of Left Sphenoid Bone with Autologous Tissue Substitute, Percutaneous  Endoscopic Approach |
|  | 0NRD4JZ | | Replacement of Left Sphenoid Bone with Synthetic Substitute, Percutaneous  Endoscopic Approach |
|  | 0NRD4KZ | | Replacement of Left Sphenoid Bone with Nonautologous Tissue Substitute,  Percutaneous Endoscopic Approach |
|  | 0NRF07Z | | Replacement of Right Ethmoid Bone with Autologous Tissue Substitute, Open Approach |
|  | 0NRF0JZ | | Replacement of Right Ethmoid Bone with Synthetic Substitute, Open Approach |
|  | 0NRF0KZ | | Replacement of Right Ethmoid Bone with Nonautologous Tissue Substitute, Open  Approach |
|  | 0NRF37Z | | Replacement of Right Ethmoid Bone with Autologous Tissue Substitute, Percutaneous  Approach |
|  | 0NRF3JZ | | Replacement of Right Ethmoid Bone with Synthetic Substitute, Percutaneous Approach |
|  | 0NRF3KZ | | Replacement of Right Ethmoid Bone with Nonautologous Tissue Substitute, |
|  |  | | Percutaneous Approach |
|  | 0NRF47Z | | Replacement of Right Ethmoid Bone with Autologous Tissue Substitute, Percutaneous Endoscopic Approach |
|  | 0NRF4JZ | | Replacement of Right Ethmoid Bone with Synthetic Substitute, Percutaneous  Endoscopic Approach |
|  | 0NRF4KZ | | Replacement of Right Ethmoid Bone with Nonautologous Tissue Substitute,  Percutaneous Endoscopic Approach |
|  | 0NRG07Z | | Replacement of Left Ethmoid Bone with Autologous Tissue Substitute, Open Approach |
|  | 0NRG0JZ | | Replacement of Left Ethmoid Bone with Synthetic Substitute, Open Approach |
|  | 0NRG0KZ | | Replacement of Left Ethmoid Bone with Nonautologous Tissue Substitute, Open Approach |
|  | 0NRG37Z | | Replacement of Left Ethmoid Bone with Autologous Tissue Substitute, Percutaneous Approach |
|  | 0NRG3JZ | | Replacement of Left Ethmoid Bone with Synthetic Substitute, Percutaneous Approach |
|  | 0NRG3KZ | | Replacement of Left Ethmoid Bone with Nonautologous Tissue Substitute,  Percutaneous Approach |
|  | 0NRG47Z | | Replacement of Left Ethmoid Bone with Autologous Tissue Substitute, Percutaneous Endoscopic Approach |
|  | 0NRG4JZ | | Replacement of Left Ethmoid Bone with Synthetic Substitute, Percutaneous Endoscopic Approach |
|  | 0NRG4KZ | | Replacement of Left Ethmoid Bone with Nonautologous Tissue Substitute, Percutaneous Endoscopic Approach |
|  | 0NRH07Z | | Replacement of Right Lacrimal Bone with Autologous Tissue Substitute, Open Approach |
|  | 0NRH0JZ | | Replacement of Right Lacrimal Bone with Synthetic Substitute, Open Approach |
|  | 0NRH0KZ | | Replacement of Right Lacrimal Bone with Nonautologous Tissue Substitute, Open Approach |
|  | 0NRH37Z | | Replacement of Right Lacrimal Bone with Autologous Tissue Substitute, Percutaneous Approach |
|  | 0NRH3JZ | | Replacement of Right Lacrimal Bone with Synthetic Substitute, Percutaneous Approach |
|  | 0NRH3KZ | | Replacement of Right Lacrimal Bone with Nonautologous Tissue Substitute,  Percutaneous Approach |
|  | 0NRH47Z | | Replacement of Right Lacrimal Bone with Autologous Tissue Substitute, Percutaneous Endoscopic Approach |
|  | 0NRH4JZ | | Replacement of Right Lacrimal Bone with Synthetic Substitute, Percutaneous Endoscopic Approach |
|  | 0NRH4KZ | | Replacement of Right Lacrimal Bone with Nonautologous Tissue Substitute, Percutaneous Endoscopic Approach |
|  | 0NRJ07Z | | Replacement of Left Lacrimal Bone with Autologous Tissue Substitute, Open Approach |
|  | 0NRJ0JZ | | Replacement of Left Lacrimal Bone with Synthetic Substitute, Open Approach |
|  | 0NRJ0KZ | | Replacement of Left Lacrimal Bone with Nonautologous Tissue Substitute, Open  Approach |
|  | 0NRJ37Z | | Replacement of Left Lacrimal Bone with Autologous Tissue Substitute, Percutaneous  Approach |
|  | 0NRJ3JZ | | Replacement of Left Lacrimal Bone with Synthetic Substitute, Percutaneous Approach |
|  | 0NRJ3KZ | | Replacement of Left Lacrimal Bone with Nonautologous Tissue Substitute, Percutaneous Approach |
|  | 0NRJ47Z | | Replacement of Left Lacrimal Bone with Autologous Tissue Substitute, Percutaneous Endoscopic Approach |
|  | 0NRJ4JZ | | Replacement of Left Lacrimal Bone with Synthetic Substitute, Percutaneous  Endoscopic Approach |
|  | 0NRJ4KZ | | Replacement of Left Lacrimal Bone with Nonautologous Tissue Substitute,  Percutaneous Endoscopic Approach |
|  | 0NRK07Z | | Replacement of Right Palatine Bone with Autologous Tissue Substitute, Open |
|  |  | | Approach |
|  | 0NRK0JZ | | Replacement of Right Palatine Bone with Synthetic Substitute, Open Approach |
|  | 0NRK0KZ | | Replacement of Right Palatine Bone with Nonautologous Tissue Substitute, Open Approach |
|  | 0NRK37Z | | Replacement of Right Palatine Bone with Autologous Tissue Substitute, Percutaneous Approach |
|  | 0NRK3JZ | | Replacement of Right Palatine Bone with Synthetic Substitute, Percutaneous Approach |
|  | 0NRK3KZ | | Replacement of Right Palatine Bone with Nonautologous Tissue Substitute,  Percutaneous Approach |
|  | 0NRK47Z | | Replacement of Right Palatine Bone with Autologous Tissue Substitute, Percutaneous Endoscopic Approach |
|  | 0NRK4JZ | | Replacement of Right Palatine Bone with Synthetic Substitute, Percutaneous Endoscopic Approach |
|  | 0NRK4KZ | | Replacement of Right Palatine Bone with Nonautologous Tissue Substitute, Percutaneous Endoscopic Approach |
|  | 0NRL07Z | | Replacement of Left Palatine Bone with Autologous Tissue Substitute, Open Approach |
|  | 0NRL0JZ | | Replacement of Left Palatine Bone with Synthetic Substitute, Open Approach |
|  | 0NRL0KZ | | Replacement of Left Palatine Bone with Nonautologous Tissue Substitute, Open  Approach |
|  | 0NRL37Z | | Replacement of Left Palatine Bone with Autologous Tissue Substitute, Percutaneous  Approach |
|  | 0NRL3JZ | | Replacement of Left Palatine Bone with Synthetic Substitute, Percutaneous Approach |
|  | 0NRL3KZ | | Replacement of Left Palatine Bone with Nonautologous Tissue Substitute, Percutaneous Approach |
|  | 0NRL47Z | | Replacement of Left Palatine Bone with Autologous Tissue Substitute, Percutaneous Endoscopic Approach |
|  | 0NRL4JZ | | Replacement of Left Palatine Bone with Synthetic Substitute, Percutaneous  Endoscopic Approach |
|  | 0NRL4KZ | | Replacement of Left Palatine Bone with Nonautologous Tissue Substitute,  Percutaneous Endoscopic Approach |
|  | 0NRM07Z | | Replacement of Right Zygomatic Bone with Autologous Tissue Substitute, Open  Approach |
|  | 0NRM0JZ | | Replacement of Right Zygomatic Bone with Synthetic Substitute, Open Approach |
|  | 0NRM0KZ | | Replacement of Right Zygomatic Bone with Nonautologous Tissue Substitute, Open  Approach |
|  | 0NRM37Z | | Replacement of Right Zygomatic Bone with Autologous Tissue Substitute,  Percutaneous Approach |
|  | 0NRM3JZ | | Replacement of Right Zygomatic Bone with Synthetic Substitute, Percutaneous  Approach |
|  | 0NRM3KZ | | Replacement of Right Zygomatic Bone with Nonautologous Tissue Substitute, Percutaneous Approach |
|  | 0NRM47Z | | Replacement of Right Zygomatic Bone with Autologous Tissue Substitute, Percutaneous Endoscopic Approach |
|  | 0NRM4JZ | | Replacement of Right Zygomatic Bone with Synthetic Substitute, Percutaneous Endoscopic Approach |
|  | 0NRM4KZ | | Replacement of Right Zygomatic Bone with Nonautologous Tissue Substitute, Percutaneous Endoscopic Approach |
|  | 0NRN07Z | | Replacement of Left Zygomatic Bone with Autologous Tissue Substitute, Open Approach |
|  | 0NRN0JZ | | Replacement of Left Zygomatic Bone with Synthetic Substitute, Open Approach |
|  | 0NRN0KZ | | Replacement of Left Zygomatic Bone with Nonautologous Tissue Substitute, Open Approach |
|  | 0NRN37Z | | Replacement of Left Zygomatic Bone with Autologous Tissue Substitute, Percutaneous Approach |
|  | 0NRN3JZ | | Replacement of Left Zygomatic Bone with Synthetic Substitute, Percutaneous  Approach |
|  | 0NRN3KZ | | Replacement of Left Zygomatic Bone with Nonautologous Tissue Substitute, Percutaneous Approach |
|  | 0NRN47Z | | Replacement of Left Zygomatic Bone with Autologous Tissue Substitute, Percutaneous Endoscopic Approach |
|  | 0NRN4JZ | | Replacement of Left Zygomatic Bone with Synthetic Substitute, Percutaneous Endoscopic Approach |
|  | 0NRN4KZ | | Replacement of Left Zygomatic Bone with Nonautologous Tissue Substitute, Percutaneous Endoscopic Approach |
|  | 0NRP0KZ | | Replacement of Right Orbit with Nonautologous Tissue Substitute, Open Approach |
|  | 0NRP3KZ | | Replacement of Right Orbit with Nonautologous Tissue Substitute, Percutaneous Approach |
|  | 0NRP4KZ | | Replacement of Right Orbit with Nonautologous Tissue Substitute, Percutaneous Endoscopic Approach |
|  | 0NRQ0KZ | | Replacement of Left Orbit with Nonautologous Tissue Substitute, Open Approach |
|  | 0NRQ3KZ | | Replacement of Left Orbit with Nonautologous Tissue Substitute, Percutaneous Approach |
|  | 0NRQ4KZ | | Replacement of Left Orbit with Nonautologous Tissue Substitute, Percutaneous Endoscopic Approach |
|  | 0NRR07Z | | Replacement of Right Maxilla with Autologous Tissue Substitute, Open Approach |
|  | 0NRR0JZ | | Replacement of Right Maxilla with Synthetic Substitute, Open Approach |
|  | 0NRR0KZ | | Replacement of Right Maxilla with Nonautologous Tissue Substitute, Open Approach |
|  | 0NRR37Z | | Replacement of Right Maxilla with Autologous Tissue Substitute, Percutaneous Approach |
|  | 0NRR3JZ | | Replacement of Right Maxilla with Synthetic Substitute, Percutaneous Approach |
|  | 0NRR3KZ | | Replacement of Right Maxilla with Nonautologous Tissue Substitute, Percutaneous  Approach |
|  | 0NRR47Z | | Replacement of Right Maxilla with Autologous Tissue Substitute, Percutaneous  Endoscopic Approach |
|  | 0NRR4JZ | | Replacement of Right Maxilla with Synthetic Substitute, Percutaneous Endoscopic  Approach |
|  | 0NRR4KZ | | Replacement of Right Maxilla with Nonautologous Tissue Substitute, Percutaneous Endoscopic Approach |
|  | 0NRS07Z | | Replacement of Left Maxilla with Autologous Tissue Substitute, Open Approach |
|  | 0NRS0JZ | | Replacement of Left Maxilla with Synthetic Substitute, Open Approach |
|  | 0NRS0KZ | | Replacement of Left Maxilla with Nonautologous Tissue Substitute, Open Approach |
|  | 0NRS37Z | | Replacement of Left Maxilla with Autologous Tissue Substitute, Percutaneous  Approach |
|  | 0NRS3JZ | | Replacement of Left Maxilla with Synthetic Substitute, Percutaneous Approach |
|  | 0NRS3KZ | | Replacement of Left Maxilla with Nonautologous Tissue Substitute, Percutaneous  Approach |
|  | 0NRS47Z | | Replacement of Left Maxilla with Autologous Tissue Substitute, Percutaneous  Endoscopic Approach |
|  | 0NRS4JZ | | Replacement of Left Maxilla with Synthetic Substitute, Percutaneous Endoscopic  Approach |
|  | 0NRS4KZ | | Replacement of Left Maxilla with Nonautologous Tissue Substitute, Percutaneous Endoscopic Approach |
|  | 0NRX07Z | | Replacement of Hyoid Bone with Autologous Tissue Substitute, Open Approach |
|  | 0NRX0JZ | | Replacement of Hyoid Bone with Synthetic Substitute, Open Approach |
|  | 0NRX0KZ | | Replacement of Hyoid Bone with Nonautologous Tissue Substitute, Open Approach |
|  | 0NRX37Z | | Replacement of Hyoid Bone with Autologous Tissue Substitute, Percutaneous  Approach |
|  | 0NRX3JZ | | Replacement of Hyoid Bone with Synthetic Substitute, Percutaneous Approach |
|  | 0NRX3KZ | | Replacement of Hyoid Bone with Nonautologous Tissue Substitute, Percutaneous Approach |
|  | 0NRX47Z | | Replacement of Hyoid Bone with Autologous Tissue Substitute, Percutaneous  Endoscopic Approach |
|  | 0NRX4JZ | | Replacement of Hyoid Bone with Synthetic Substitute, Percutaneous Endoscopic  Approach |
|  | 0NRX4KZ | | Replacement of Hyoid Bone with Nonautologous Tissue Substitute, Percutaneous  Endoscopic Approach |
|  | 0NUC07Z | | Supplement Right Sphenoid Bone with Autologous Tissue Substitute, Open Approach |
|  | 0NUC0JZ | | Supplement Right Sphenoid Bone with Synthetic Substitute, Open Approach |
|  | 0NUC0KZ | | Supplement Right Sphenoid Bone with Nonautologous Tissue Substitute, Open Approach |
|  | 0NUC37Z | | Supplement Right Sphenoid Bone with Autologous Tissue Substitute, Percutaneous Approach |
|  | 0NUC3JZ | | Supplement Right Sphenoid Bone with Synthetic Substitute, Percutaneous Approach |
|  | 0NUC3KZ | | Supplement Right Sphenoid Bone with Nonautologous Tissue Substitute, Percutaneous Approach |
|  | 0NUC47Z | | Supplement Right Sphenoid Bone with Autologous Tissue Substitute, Percutaneous Endoscopic Approach |
|  | 0NUC4JZ | | Supplement Right Sphenoid Bone with Synthetic Substitute, Percutaneous Endoscopic Approach |
|  | 0NUC4KZ | | Supplement Right Sphenoid Bone with Nonautologous Tissue Substitute, Percutaneous Endoscopic Approach |
|  | 0NUD07Z | | Supplement Left Sphenoid Bone with Autologous Tissue Substitute, Open Approach |
|  | 0NUD0JZ | | Supplement Left Sphenoid Bone with Synthetic Substitute, Open Approach |
|  | 0NUD0KZ | | Supplement Left Sphenoid Bone with Nonautologous Tissue Substitute, Open  Approach |
|  | 0NUD37Z | | Supplement Left Sphenoid Bone with Autologous Tissue Substitute, Percutaneous  Approach |
|  | 0NUD3JZ | | Supplement Left Sphenoid Bone with Synthetic Substitute, Percutaneous Approach |
|  | 0NUD3KZ | | Supplement Left Sphenoid Bone with Nonautologous Tissue Substitute, Percutaneous Approach |
|  | 0NUD47Z | | Supplement Left Sphenoid Bone with Autologous Tissue Substitute, Percutaneous  Endoscopic Approach |
|  | 0NUD4JZ | | Supplement Left Sphenoid Bone with Synthetic Substitute, Percutaneous Endoscopic  Approach |
|  | 0NUD4KZ | | Supplement Left Sphenoid Bone with Nonautologous Tissue Substitute, Percutaneous  Endoscopic Approach |
|  | 0NUF07Z | | Supplement Right Ethmoid Bone with Autologous Tissue Substitute, Open Approach |
|  | 0NUF0JZ | | Supplement Right Ethmoid Bone with Synthetic Substitute, Open Approach |
|  | 0NUF0KZ | | Supplement Right Ethmoid Bone with Nonautologous Tissue Substitute, Open Approach |
|  | 0NUF37Z | | Supplement Right Ethmoid Bone with Autologous Tissue Substitute, Percutaneous Approach |
|  | 0NUF3JZ | | Supplement Right Ethmoid Bone with Synthetic Substitute, Percutaneous Approach |
|  | 0NUF3KZ | | Supplement Right Ethmoid Bone with Nonautologous Tissue Substitute, Percutaneous Approach |
|  | 0NUF47Z | | Supplement Right Ethmoid Bone with Autologous Tissue Substitute, Percutaneous Endoscopic Approach |
|  | 0NUF4JZ | | Supplement Right Ethmoid Bone with Synthetic Substitute, Percutaneous Endoscopic Approach |
|  | 0NUF4KZ | | Supplement Right Ethmoid Bone with Nonautologous Tissue Substitute, Percutaneous Endoscopic Approach |
| 0NUG07Z | | Supplement Left Ethmoid Bone with Autologous Tissue Substitute, Open Approach | |
| 0NUG0JZ | | Supplement Left Ethmoid Bone with Synthetic Substitute, Open Approach | |
| 0NUG0KZ | | Supplement Left Ethmoid Bone with Nonautologous Tissue Substitute, Open Approach | |
| 0NUG37Z | | Supplement Left Ethmoid Bone with Autologous Tissue Substitute, Percutaneous  Approach | |
| 0NUG3JZ | | Supplement Left Ethmoid Bone with Synthetic Substitute, Percutaneous Approach | |
| 0NUG3KZ | | Supplement Left Ethmoid Bone with Nonautologous Tissue Substitute, Percutaneous Approach | |
| 0NUG47Z | | Supplement Left Ethmoid Bone with Autologous Tissue Substitute, Percutaneous Endoscopic Approach | |
| 0NUG4JZ | | Supplement Left Ethmoid Bone with Synthetic Substitute, Percutaneous Endoscopic  Approach | |
| 0NUG4KZ | | Supplement Left Ethmoid Bone with Nonautologous Tissue Substitute, Percutaneous  Endoscopic Approach | |
| 0NUH07Z | | Supplement Right Lacrimal Bone with Autologous Tissue Substitute, Open Approach | |
| 0NUH0JZ | | Supplement Right Lacrimal Bone with Synthetic Substitute, Open Approach | |
| 0NUH0KZ | | Supplement Right Lacrimal Bone with Nonautologous Tissue Substitute, Open Approach | |
| 0NUH37Z | | Supplement Right Lacrimal Bone with Autologous Tissue Substitute, Percutaneous Approach | |
| 0NUH3JZ | | Supplement Right Lacrimal Bone with Synthetic Substitute, Percutaneous Approach | |
| 0NUH3KZ | | Supplement Right Lacrimal Bone with Nonautologous Tissue Substitute, Percutaneous  Approach | |
| 0NUH47Z | | Supplement Right Lacrimal Bone with Autologous Tissue Substitute, Percutaneous Endoscopic Approach | |
| 0NUH4JZ | | Supplement Right Lacrimal Bone with Synthetic Substitute, Percutaneous Endoscopic Approach | |
| 0NUH4KZ | | Supplement Right Lacrimal Bone with Nonautologous Tissue Substitute, Percutaneous Endoscopic Approach | |
| 0NUJ07Z | | Supplement Left Lacrimal Bone with Autologous Tissue Substitute, Open Approach | |
| 0NUJ0JZ | | Supplement Left Lacrimal Bone with Synthetic Substitute, Open Approach | |
| 0NUJ0KZ | | Supplement Left Lacrimal Bone with Nonautologous Tissue Substitute, Open Approach | |
| 0NUJ37Z | | Supplement Left Lacrimal Bone with Autologous Tissue Substitute, Percutaneous Approach | |
| 0NUJ3JZ | | Supplement Left Lacrimal Bone with Synthetic Substitute, Percutaneous Approach | |
| 0NUJ3KZ | | Supplement Left Lacrimal Bone with Nonautologous Tissue Substitute, Percutaneous  Approach | |
| 0NUJ47Z | | Supplement Left Lacrimal Bone with Autologous Tissue Substitute, Percutaneous Endoscopic Approach | |
| 0NUJ4JZ | | Supplement Left Lacrimal Bone with Synthetic Substitute, Percutaneous Endoscopic Approach | |
| 0NUJ4KZ | | Supplement Left Lacrimal Bone with Nonautologous Tissue Substitute, Percutaneous Endoscopic Approach | |
| 0NUK07Z | | Supplement Right Palatine Bone with Autologous Tissue Substitute, Open Approach | |
| 0NUK0JZ | | Supplement Right Palatine Bone with Synthetic Substitute, Open Approach | |
| 0NUK0KZ | | Supplement Right Palatine Bone with Nonautologous Tissue Substitute, Open Approach | |
| 0NUK37Z | | Supplement Right Palatine Bone with Autologous Tissue Substitute, Percutaneous  Approach | |
| 0NUK3JZ | | Supplement Right Palatine Bone with Synthetic Substitute, Percutaneous Approach | |
| 0NUK3KZ | | Supplement Right Palatine Bone with Nonautologous Tissue Substitute, Percutaneous Approach | |
| 0NUK47Z | | Supplement Right Palatine Bone with Autologous Tissue Substitute, Percutaneous | |

|  | Endoscopic Approach |
| --- | --- |
| 0NUK4JZ | Supplement Right Palatine Bone with Synthetic Substitute, Percutaneous Endoscopic Approach |
| 0NUK4KZ | Supplement Right Palatine Bone with Nonautologous Tissue Substitute, Percutaneous  Endoscopic Approach |
| 0NUL07Z | Supplement Left Palatine Bone with Autologous Tissue Substitute, Open Approach |
| 0NUL0JZ | Supplement Left Palatine Bone with Synthetic Substitute, Open Approach |
| 0NUL0KZ | Supplement Left Palatine Bone with Nonautologous Tissue Substitute, Open Approach |
| 0NUL37Z | Supplement Left Palatine Bone with Autologous Tissue Substitute, Percutaneous Approach |
| 0NUL3JZ | Supplement Left Palatine Bone with Synthetic Substitute, Percutaneous Approach |
| 0NUL3KZ | Supplement Left Palatine Bone with Nonautologous Tissue Substitute, Percutaneous Approach |
| 0NUL47Z | Supplement Left Palatine Bone with Autologous Tissue Substitute, Percutaneous Endoscopic Approach |
| 0NUL4JZ | Supplement Left Palatine Bone with Synthetic Substitute, Percutaneous Endoscopic Approach |
| 0NUL4KZ | Supplement Left Palatine Bone with Nonautologous Tissue Substitute, Percutaneous  Endoscopic Approach |
| 0NUM07Z | Supplement Right Zygomatic Bone with Autologous Tissue Substitute, Open Approach |
| 0NUM0JZ | Supplement Right Zygomatic Bone with Synthetic Substitute, Open Approach |
| 0NUM0KZ | Supplement Right Zygomatic Bone with Nonautologous Tissue Substitute, Open  Approach |
| 0NUM37Z | Supplement Right Zygomatic Bone with Autologous Tissue Substitute, Percutaneous Approach |
| 0NUM3JZ | Supplement Right Zygomatic Bone with Synthetic Substitute, Percutaneous Approach |
| 0NUM3KZ | Supplement Right Zygomatic Bone with Nonautologous Tissue Substitute,  Percutaneous Approach |
| 0NUM47Z | Supplement Right Zygomatic Bone with Autologous Tissue Substitute, Percutaneous  Endoscopic Approach |
| 0NUM4JZ | Supplement Right Zygomatic Bone with Synthetic Substitute, Percutaneous Endoscopic Approach |
| 0NUM4KZ | Supplement Right Zygomatic Bone with Nonautologous Tissue Substitute, Percutaneous Endoscopic Approach |
| 0NUN07Z | Supplement Left Zygomatic Bone with Autologous Tissue Substitute, Open Approach |
| 0NUN0JZ | Supplement Left Zygomatic Bone with Synthetic Substitute, Open Approach |
| 0NUN0KZ | Supplement Left Zygomatic Bone with Nonautologous Tissue Substitute, Open Approach |
| 0NUN37Z | Supplement Left Zygomatic Bone with Autologous Tissue Substitute, Percutaneous Approach |
| 0NUN3JZ | Supplement Left Zygomatic Bone with Synthetic Substitute, Percutaneous Approach |
| 0NUN3KZ | Supplement Left Zygomatic Bone with Nonautologous Tissue Substitute, Percutaneous Approach |
| 0NUN47Z | Supplement Left Zygomatic Bone with Autologous Tissue Substitute, Percutaneous Endoscopic Approach |
| 0NUN4JZ | Supplement Left Zygomatic Bone with Synthetic Substitute, Percutaneous Endoscopic Approach |
| 0NUN4KZ | Supplement Left Zygomatic Bone with Nonautologous Tissue Substitute, Percutaneous  Endoscopic Approach |
| 0NUP07Z | Supplement Right Orbit with Autologous Tissue Substitute, Open Approach |
| 0NUP0KZ | Supplement Right Orbit with Nonautologous Tissue Substitute, Open Approach |
| 0NUP37Z | Supplement Right Orbit with Autologous Tissue Substitute, Percutaneous Approach |
| 0NUP3KZ | Supplement Right Orbit with Nonautologous Tissue Substitute, Percutaneous Approach |

| 0NUP47Z | Supplement Right Orbit with Autologous Tissue Substitute, Percutaneous Endoscopic  Approach |
| --- | --- |
| 0NUP4KZ | Supplement Right Orbit with Nonautologous Tissue Substitute, Percutaneous Endoscopic Approach |
| 0NUQ07Z | Supplement Left Orbit with Autologous Tissue Substitute, Open Approach |
| 0NUQ0KZ | Supplement Left Orbit with Nonautologous Tissue Substitute, Open Approach |
| 0NUQ37Z | Supplement Left Orbit with Autologous Tissue Substitute, Percutaneous Approach |
| 0NUQ3KZ | Supplement Left Orbit with Nonautologous Tissue Substitute, Percutaneous Approach |
| 0NUQ47Z | Supplement Left Orbit with Autologous Tissue Substitute, Percutaneous Endoscopic Approach |
| 0NUQ4KZ | Supplement Left Orbit with Nonautologous Tissue Substitute, Percutaneous  Endoscopic Approach |
| 0NUR07Z | Supplement Right Maxilla with Autologous Tissue Substitute, Open Approach |
| 0NUR0JZ | Supplement Right Maxilla with Synthetic Substitute, Open Approach |
| 0NUR0KZ | Supplement Right Maxilla with Nonautologous Tissue Substitute, Open Approach |
| 0NUR37Z | Supplement Right Maxilla with Autologous Tissue Substitute, Percutaneous Approach |
| 0NUR3JZ | Supplement Right Maxilla with Synthetic Substitute, Percutaneous Approach |
| 0NUR3KZ | Supplement Right Maxilla with Nonautologous Tissue Substitute, Percutaneous  Approach |
| 0NUR47Z | Supplement Right Maxilla with Autologous Tissue Substitute, Percutaneous  Endoscopic Approach |
| 0NUR4JZ | Supplement Right Maxilla with Synthetic Substitute, Percutaneous Endoscopic  Approach |
| 0NUR4KZ | Supplement Right Maxilla with Nonautologous Tissue Substitute, Percutaneous Endoscopic Approach |
| 0NUS07Z | Supplement Left Maxilla with Autologous Tissue Substitute, Open Approach |
| 0NUS0JZ | Supplement Left Maxilla with Synthetic Substitute, Open Approach |
| 0NUS0KZ | Supplement Left Maxilla with Nonautologous Tissue Substitute, Open Approach |
| 0NUS37Z | Supplement Left Maxilla with Autologous Tissue Substitute, Percutaneous Approach |
| 0NUS3JZ | Supplement Left Maxilla with Synthetic Substitute, Percutaneous Approach |
| 0NUS3KZ | Supplement Left Maxilla with Nonautologous Tissue Substitute, Percutaneous Approach |
| 0NUS47Z | Supplement Left Maxilla with Autologous Tissue Substitute, Percutaneous Endoscopic Approach |
| 0NUS4JZ | Supplement Left Maxilla with Synthetic Substitute, Percutaneous Endoscopic Approach |
| 0NUS4KZ | Supplement Left Maxilla with Nonautologous Tissue Substitute, Percutaneous  Endoscopic Approach |
| 0NUX07Z | Supplement Hyoid Bone with Autologous Tissue Substitute, Open Approach |
| 0NUX0JZ | Supplement Hyoid Bone with Synthetic Substitute, Open Approach |
| 0NUX0KZ | Supplement Hyoid Bone with Nonautologous Tissue Substitute, Open Approach |
| 0NUX37Z | Supplement Hyoid Bone with Autologous Tissue Substitute, Percutaneous Approach |
| 0NUX3JZ | Supplement Hyoid Bone with Synthetic Substitute, Percutaneous Approach |
| 0NUX3KZ | Supplement Hyoid Bone with Nonautologous Tissue Substitute, Percutaneous Approach |
| 0NUX47Z | Supplement Hyoid Bone with Autologous Tissue Substitute, Percutaneous Endoscopic Approach |
| 0NUX4JZ | Supplement Hyoid Bone with Synthetic Substitute, Percutaneous Endoscopic Approach |
| 0NUX4KZ | Supplement Hyoid Bone with Nonautologous Tissue Substitute, Percutaneous  Endoscopic Approach |
| 0RNC0ZZ | Release Right Temporomandibular Joint, Open Approach |
| 0RNC3ZZ | Release Right Temporomandibular Joint, Percutaneous Approach |
| 0RNC4ZZ | Release Right Temporomandibular Joint, Percutaneous Endoscopic Approach |
| 0RND0ZZ | Release Left Temporomandibular Joint, Open Approach |

| 0RND3ZZ | Release Left Temporomandibular Joint, Percutaneous Approach |
| --- | --- |
| 0RND4ZZ | Release Left Temporomandibular Joint, Percutaneous Endoscopic Approach |
| 0W0407Z | Alteration of Upper Jaw with Autologous Tissue Substitute, Open Approach |
| 0W040JZ | Alteration of Upper Jaw with Synthetic Substitute, Open Approach |
| 0W040KZ | Alteration of Upper Jaw with Nonautologous Tissue Substitute, Open Approach |
| 0W040ZZ | Alteration of Upper Jaw, Open Approach |
| 0W0437Z | Alteration of Upper Jaw with Autologous Tissue Substitute, Percutaneous Approach |
| 0W043JZ | Alteration of Upper Jaw with Synthetic Substitute, Percutaneous Approach |
| 0W043KZ | Alteration of Upper Jaw with Nonautologous Tissue Substitute, Percutaneous Approach |
| 0W043ZZ | Alteration of Upper Jaw, Percutaneous Approach |
| 0W0447Z | Alteration of Upper Jaw with Autologous Tissue Substitute, Percutaneous Endoscopic  Approach |
| 0W044JZ | Alteration of Upper Jaw with Synthetic Substitute, Percutaneous Endoscopic Approach |
| 0W044KZ | Alteration of Upper Jaw with Nonautologous Tissue Substitute, Percutaneous Endoscopic Approach |
| 0W044ZZ | Alteration of Upper Jaw, Percutaneous Endoscopic Approach |
| 0W0507Z | Alteration of Lower Jaw with Autologous Tissue Substitute, Open Approach |
| 0W050JZ | Alteration of Lower Jaw with Synthetic Substitute, Open Approach |
| 0W050KZ | Alteration of Lower Jaw with Nonautologous Tissue Substitute, Open Approach |
| 0W050ZZ | Alteration of Lower Jaw, Open Approach |
| 0W0537Z | Alteration of Lower Jaw with Autologous Tissue Substitute, Percutaneous Approach |
| 0W053JZ | Alteration of Lower Jaw with Synthetic Substitute, Percutaneous Approach |
| 0W053KZ | Alteration of Lower Jaw with Nonautologous Tissue Substitute, Percutaneous Approach |
| 0W053ZZ | Alteration of Lower Jaw, Percutaneous Approach |
| 0W0547Z | Alteration of Lower Jaw with Autologous Tissue Substitute, Percutaneous Endoscopic  Approach |
| 0W054JZ | Alteration of Lower Jaw with Synthetic Substitute, Percutaneous Endoscopic Approach |
| 0W054KZ | Alteration of Lower Jaw with Nonautologous Tissue Substitute, Percutaneous Endoscopic Approach |
| 0W054ZZ | Alteration of Lower Jaw, Percutaneous Endoscopic Approach |
| 0WU407Z | Supplement Upper Jaw with Autologous Tissue Substitute, Open Approach |
| 0WU40JZ | Supplement Upper Jaw with Synthetic Substitute, Open Approach |
| 0WU40KZ | Supplement Upper Jaw with Nonautologous Tissue Substitute, Open Approach |
| 0WU447Z | Supplement Upper Jaw with Autologous Tissue Substitute, Percutaneous Endoscopic Approach |
| 0WU44JZ | Supplement Upper Jaw with Synthetic Substitute, Percutaneous Endoscopic Approach |
| 0WU44KZ | Supplement Upper Jaw with Nonautologous Tissue Substitute, Percutaneous  Endoscopic Approach |
| 0WU507Z | Supplement Lower Jaw with Autologous Tissue Substitute, Open Approach |
| 0WU50JZ | Supplement Lower Jaw with Synthetic Substitute, Open Approach |
| 0WU50KZ | Supplement Lower Jaw with Nonautologous Tissue Substitute, Open Approach |
| 0WU547Z | Supplement Lower Jaw with Autologous Tissue Substitute, Percutaneous Endoscopic  Approach |
| 0WU54JZ | Supplement Lower Jaw with Synthetic Substitute, Percutaneous Endoscopic Approach |
| 0WU54KZ | Supplement Lower Jaw with Nonautologous Tissue Substitute, Percutaneous Endoscopic Approach |

**Female to Male Surgery**

| **ICD-10-PCS**  **procedure codes:** | **Code Description** |
| --- | --- |
| 0VTC0ZZ | Resection of Bilateral Testes, Open Approach |

| 0H0T0ZZ | Alteration of Right Breast, Open Approach |
| --- | --- |
| 0H0T3ZZ | Alteration of Right Breast, Percutaneous Approach |
| 0H0TXZZ | Alteration of Right Breast, External Approach |
| 0H0U0ZZ | Alteration of Left Breast, Open Approach |
| 0H0U3ZZ | Alteration of Left Breast, Percutaneous Approach |
| 0H0UXZZ | Alteration of Left Breast, External Approach |
| 0H0V07Z | Alteration of Bilateral Breast with Autologous Tissue Substitute, Open Approach |
| 0H0V0JZ | Alteration of Bilateral Breast with Synthetic Substitute, Open Approach |
| 0H0V0KZ | Alteration of Bilateral Breast with Nonautologous Tissue Substitute, Open Approach |
| 0H0V0ZZ | Alteration of Bilateral Breast, Open Approach |
| 0H0V37Z | Alteration of Bilateral Breast with Autologous Tissue Substitute, Percutaneous  Approach |
| 0H0V3JZ | Alteration of Bilateral Breast with Synthetic Substitute, Percutaneous Approach |
| 0H0V3KZ | Alteration of Bilateral Breast with Nonautologous Tissue Substitute, Percutaneous Approach |
| 0H0V3ZZ | Alteration of Bilateral Breast, Percutaneous Approach |
| 0H0VXZZ | Alteration of Bilateral Breast, External Approach |
| 0HMTXZZ | Reattachment of Right Breast, External Approach |
| 0HMUXZZ | Reattachment of Left Breast, External Approach |
| 0HMVXZZ | Reattachment of Bilateral Breast, External Approach |
| 0HMWXZZ | Reattachment of Right Nipple, External Approach |
| 0HMXXZZ | Reattachment of Left Nipple, External Approach |
| 0HNT0ZZ | Release Right Breast, Open Approach |
| 0HNT3ZZ | Release Right Breast, Percutaneous Approach |
| 0HNT7ZZ | Release Right Breast, Via Natural or Artificial Opening |
| 0HNT8ZZ | Release Right Breast, Via Natural or Artificial Opening Endoscopic |
| 0HNTXZZ | Release Right Breast, External Approach |
| 0HNU0ZZ | Release Left Breast, Open Approach |
| 0HNU3ZZ | Release Left Breast, Percutaneous Approach |
| 0HNU7ZZ | Release Left Breast, Via Natural or Artificial Opening |
| 0HNU8ZZ | Release Left Breast, Via Natural or Artificial Opening Endoscopic |
| 0HNUXZZ | Release Left Breast, External Approach |
| 0HNV0ZZ | Release Bilateral Breast, Open Approach |
| 0HNV3ZZ | Release Bilateral Breast, Percutaneous Approach |
| 0HNV7ZZ | Release Bilateral Breast, Via Natural or Artificial Opening |
| 0HNV8ZZ | Release Bilateral Breast, Via Natural or Artificial Opening Endoscopic |
| 0HNVXZZ | Release Bilateral Breast, External Approach |
| 0HNW0ZZ | Release Right Nipple, Open Approach |
| 0HNW3ZZ | Release Right Nipple, Percutaneous Approach |
| 0HNW7ZZ | Release Right Nipple, Via Natural or Artificial Opening |
| 0HNW8ZZ | Release Right Nipple, Via Natural or Artificial Opening Endoscopic |
| 0HNWXZZ | Release Right Nipple, External Approach |
| 0HNX0ZZ | Release Left Nipple, Open Approach |
| 0HNX3ZZ | Release Left Nipple, Percutaneous Approach |
| 0HNX7ZZ | Release Left Nipple, Via Natural or Artificial Opening |
| 0HNX8ZZ | Release Left Nipple, Via Natural or Artificial Opening Endoscopic |
| 0HNXXZZ | Release Left Nipple, External Approach |
| 0HQT0ZZ | Repair Right Breast, Open Approach |
| 0HQT3ZZ | Repair Right Breast, Percutaneous Approach |
| 0HQT7ZZ | Repair Right Breast, Via Natural or Artificial Opening |
| 0HQT8ZZ | Repair Right Breast, Via Natural or Artificial Opening Endoscopic |

| 0HQTXZZ | Repair Right Breast, External Approach |
| --- | --- |
| 0HQU0ZZ | Repair Left Breast, Open Approach |
| 0HQU3ZZ | Repair Left Breast, Percutaneous Approach |
| 0HQU7ZZ | Repair Left Breast, Via Natural or Artificial Opening |
| 0HQU8ZZ | Repair Left Breast, Via Natural or Artificial Opening Endoscopic |
| 0HQUXZZ | Repair Left Breast, External Approach |
| 0HQV0ZZ | Repair Bilateral Breast, Open Approach |
| 0HQV3ZZ | Repair Bilateral Breast, Percutaneous Approach |
| 0HQV7ZZ | Repair Bilateral Breast, Via Natural or Artificial Opening |
| 0HQV8ZZ | Repair Bilateral Breast, Via Natural or Artificial Opening Endoscopic |
| 0HQVXZZ | Repair Bilateral Breast, External Approach |
| 0HQW0ZZ | Repair Right Nipple, Open Approach |
| 0HQW3ZZ | Repair Right Nipple, Percutaneous Approach |
| 0HQW7ZZ | Repair Right Nipple, Via Natural or Artificial Opening |
| 0HQW8ZZ | Repair Right Nipple, Via Natural or Artificial Opening Endoscopic |
| 0HQWXZZ | Repair Right Nipple, External Approach |
| 0HQX0ZZ | Repair Left Nipple, Open Approach |
| 0HQX3ZZ | Repair Left Nipple, Percutaneous Approach |
| 0HQX7ZZ | Repair Left Nipple, Via Natural or Artificial Opening |
| 0HQX8ZZ | Repair Left Nipple, Via Natural or Artificial Opening Endoscopic |
| 0HQXXZZ | Repair Left Nipple, External Approach |
| 0HQY0ZZ | Repair Supernumerary Breast, Open Approach |
| 0HQY3ZZ | Repair Supernumerary Breast, Percutaneous Approach |
| 0HQY7ZZ | Repair Supernumerary Breast, Via Natural or Artificial Opening |
| 0HQY8ZZ | Repair Supernumerary Breast, Via Natural or Artificial Opening Endoscopic |
| 0HQYXZZ | Repair Supernumerary Breast, External Approach |
| 0HRT07Z | Replacement of Right Breast with Autologous Tissue Substitute, Open Approach |
| 0HRT0KZ | Replacement of Right Breast with Nonautologous Tissue Substitute, Open Approach |
| 0HRT37Z | Replacement of Right Breast with Autologous Tissue Substitute, Percutaneous Approach |
| 0HRT3KZ | Replacement of Right Breast with Nonautologous Tissue Substitute, Percutaneous Approach |
| 0HRTXJZ | Replacement of Right Breast with Synthetic Substitute, External Approach |
| 0HRU07Z | Replacement of Left Breast with Autologous Tissue Substitute, Open Approach |
| 0HRU0KZ | Replacement of Left Breast with Nonautologous Tissue Substitute, Open Approach |
| 0HRU37Z | Replacement of Left Breast with Autologous Tissue Substitute, Percutaneous Approach |
| 0HRU3KZ | Replacement of Left Breast with Nonautologous Tissue Substitute, Percutaneous  Approach |
| 0HRUXJZ | Replacement of Left Breast with Synthetic Substitute, External Approach |
| 0HRV07Z | Replacement of Bilateral Breast with Autologous Tissue Substitute, Open Approach |
| 0HRV0KZ | Replacement of Bilateral Breast with Nonautologous Tissue Substitute, Open Approach |
| 0HRV37Z | Replacement of Bilateral Breast with Autologous Tissue Substitute, Percutaneous  Approach |
| 0HRV3KZ | Replacement of Bilateral Breast with Nonautologous Tissue Substitute, Percutaneous  Approach |
| 0HRVXJZ | Replacement of Bilateral Breast with Synthetic Substitute, External Approach |
| 0HRW07Z | Replacement of Right Nipple with Autologous Tissue Substitute, Open Approach |
| 0HRW0JZ | Replacement of Right Nipple with Synthetic Substitute, Open Approach |
| 0HRW0KZ | Replacement of Right Nipple with Nonautologous Tissue Substitute, Open Approach |
| 0HRW37Z | Replacement of Right Nipple with Autologous Tissue Substitute, Percutaneous Approach |

| 0HRW3JZ | Replacement of Right Nipple with Synthetic Substitute, Percutaneous Approach |
| --- | --- |
| 0HRW3KZ | Replacement of Right Nipple with Nonautologous Tissue Substitute, Percutaneous Approach |
| 0HRWX7Z | Replacement of Right Nipple with Autologous Tissue Substitute, External Approach |
| 0HRWXJZ | Replacement of Right Nipple with Synthetic Substitute, External Approach |
| 0HRWXKZ | Replacement of Right Nipple with Nonautologous Tissue Substitute, External Approach |
| 0HRX07Z | Replacement of Left Nipple with Autologous Tissue Substitute, Open Approach |
| 0HRX0JZ | Replacement of Left Nipple with Synthetic Substitute, Open Approach |
| 0HRX0KZ | Replacement of Left Nipple with Nonautologous Tissue Substitute, Open Approach |
| 0HRX37Z | Replacement of Left Nipple with Autologous Tissue Substitute, Percutaneous Approach |
| 0HRX3JZ | Replacement of Left Nipple with Synthetic Substitute, Percutaneous Approach |
| 0HRX3KZ | Replacement of Left Nipple with Nonautologous Tissue Substitute, Percutaneous Approach |
| 0HRXX7Z | Replacement of Left Nipple with Autologous Tissue Substitute, External Approach |
| 0HRXXJZ | Replacement of Left Nipple with Synthetic Substitute, External Approach |
| 0HRXXKZ | Replacement of Left Nipple with Nonautologous Tissue Substitute, External Approach |
| 0HUT07Z | Supplement Right Breast with Autologous Tissue Substitute, Open Approach |
| 0HUT0JZ | Supplement Right Breast with Synthetic Substitute, Open Approach |
| 0HUT0KZ | Supplement Right Breast with Nonautologous Tissue Substitute, Open Approach |
| 0HUT37Z | Supplement Right Breast with Autologous Tissue Substitute, Percutaneous Approach |
| 0HUT3JZ | Supplement Right Breast with Synthetic Substitute, Percutaneous Approach |
| 0HUT3KZ | Supplement Right Breast with Nonautologous Tissue Substitute, Percutaneous Approach |
| 0HUT77Z | Supplement Right Breast with Autologous Tissue Substitute, Via Natural or Artificial Opening |
| 0HUT7JZ | Supplement Right Breast with Synthetic Substitute, Via Natural or Artificial Opening |
| 0HUT7KZ | Supplement Right Breast with Nonautologous Tissue Substitute, Via Natural or Artificial  Opening |
| 0HUT87Z | Supplement Right Breast with Autologous Tissue Substitute, Via Natural or Artificial Opening Endoscopic |
| 0HUT8JZ | Supplement Right Breast with Synthetic Substitute, Via Natural or Artificial Opening Endoscopic |
| 0HUT8KZ | Supplement Right Breast with Nonautologous Tissue Substitute, Via Natural or Artificial Opening Endoscopic |
| 0HUTX7Z | Supplement Right Breast with Autologous Tissue Substitute, External Approach |
| 0HUTXJZ | Supplement Right Breast with Synthetic Substitute, External Approach |
| 0HUTXKZ | Supplement Right Breast with Nonautologous Tissue Substitute, External Approach |
| 0HUU07Z | Supplement Left Breast with Autologous Tissue Substitute, Open Approach |
| 0HUU0JZ | Supplement Left Breast with Synthetic Substitute, Open Approach |
| 0HUU0KZ | Supplement Left Breast with Nonautologous Tissue Substitute, Open Approach |
| 0HUU37Z | Supplement Left Breast with Autologous Tissue Substitute, Percutaneous Approach |
| 0HUU3JZ | Supplement Left Breast with Synthetic Substitute, Percutaneous Approach |
| 0HUU3KZ | Supplement Left Breast with Nonautologous Tissue Substitute, Percutaneous Approach |
| 0HUU77Z | Supplement Left Breast with Autologous Tissue Substitute, Via Natural or Artificial  Opening |
| 0HUU7JZ | Supplement Left Breast with Synthetic Substitute, Via Natural or Artificial Opening |
| 0HUU7KZ | Supplement Left Breast with Nonautologous Tissue Substitute, Via Natural or Artificial Opening |
| 0HUU87Z | Supplement Left Breast with Autologous Tissue Substitute, Via Natural or Artificial Opening Endoscopic |
| 0HUU8JZ | Supplement Left Breast with Synthetic Substitute, Via Natural or Artificial Opening  Endoscopic |

| 0HUU8KZ | Supplement Left Breast with Nonautologous Tissue Substitute, Via Natural or Artificial  Opening Endoscopic |
| --- | --- |
| 0HUUX7Z | Supplement Left Breast with Autologous Tissue Substitute, External Approach |
| 0HUUXJZ | Supplement Left Breast with Synthetic Substitute, External Approach |
| 0HUUXKZ | Supplement Left Breast with Nonautologous Tissue Substitute, External Approach |
| 0HUV07Z | Supplement Bilateral Breast with Autologous Tissue Substitute, Open Approach |
| 0HUV0JZ | Supplement Bilateral Breast with Synthetic Substitute, Open Approach |
| 0HUV0KZ | Supplement Bilateral Breast with Nonautologous Tissue Substitute, Open Approach |
| 0HUV37Z | Supplement Bilateral Breast with Autologous Tissue Substitute, Percutaneous Approach |
| 0HUV3JZ | Supplement Bilateral Breast with Synthetic Substitute, Percutaneous Approach |
| 0HUV3KZ | Supplement Bilateral Breast with Nonautologous Tissue Substitute, Percutaneous Approach |
| 0HUV77Z | Supplement Bilateral Breast with Autologous Tissue Substitute, Via Natural or Artificial Opening |
| 0HUV7JZ | Supplement Bilateral Breast with Synthetic Substitute, Via Natural or Artificial Opening |
| 0HUV7KZ | Supplement Bilateral Breast with Nonautologous Tissue Substitute, Via Natural or Artificial Opening |
| 0HUV87Z | Supplement Bilateral Breast with Autologous Tissue Substitute, Via Natural or Artificial Opening Endoscopic |
| 0HUV8JZ | Supplement Bilateral Breast with Synthetic Substitute, Via Natural or Artificial Opening Endoscopic |
| 0HUV8KZ | Supplement Bilateral Breast with Nonautologous Tissue Substitute, Via Natural or Artificial Opening Endoscopic |
| 0HUVX7Z | Supplement Bilateral Breast with Autologous Tissue Substitute, External Approach |
| 0HUVXJZ | Supplement Bilateral Breast with Synthetic Substitute, External Approach |
| 0HUVXKZ | Supplement Bilateral Breast with Nonautologous Tissue Substitute, External Approach |
| 0HUW07Z | Supplement Right Nipple with Autologous Tissue Substitute, Open Approach |
| 0HUW0JZ | Supplement Right Nipple with Synthetic Substitute, Open Approach |
| 0HUW0KZ | Supplement Right Nipple with Nonautologous Tissue Substitute, Open Approach |
| 0HUW37Z | Supplement Right Nipple with Autologous Tissue Substitute, Percutaneous Approach |
| 0HUW3JZ | Supplement Right Nipple with Synthetic Substitute, Percutaneous Approach |
| 0HUW3KZ | Supplement Right Nipple with Nonautologous Tissue Substitute, Percutaneous Approach |
| 0HUW77Z | Supplement Right Nipple with Autologous Tissue Substitute, Via Natural or Artificial Opening |
| 0HUW7JZ | Supplement Right Nipple with Synthetic Substitute, Via Natural or Artificial Opening |
| 0HUW7KZ | Supplement Right Nipple with Nonautologous Tissue Substitute, Via Natural or Artificial Opening |
| 0HUW87Z | Supplement Right Nipple with Autologous Tissue Substitute, Via Natural or Artificial Opening Endoscopic |
| 0HUW8JZ | Supplement Right Nipple with Synthetic Substitute, Via Natural or Artificial Opening Endoscopic |
| 0HUW8KZ | Supplement Right Nipple with Nonautologous Tissue Substitute, Via Natural or Artificial Opening Endoscopic |
| 0HUWX7Z | Supplement Right Nipple with Autologous Tissue Substitute, External Approach |
| 0HUWXJZ | Supplement Right Nipple with Synthetic Substitute, External Approach |
| 0HUWXKZ | Supplement Right Nipple with Nonautologous Tissue Substitute, External Approach |
| 0HUX07Z | Supplement Left Nipple with Autologous Tissue Substitute, Open Approach |
| 0HUX0JZ | Supplement Left Nipple with Synthetic Substitute, Open Approach |
| 0HUX0KZ | Supplement Left Nipple with Nonautologous Tissue Substitute, Open Approach |
| 0HUX37Z | Supplement Left Nipple with Autologous Tissue Substitute, Percutaneous Approach |
| 0HUX3JZ | Supplement Left Nipple with Synthetic Substitute, Percutaneous Approach |

| 0HUX3KZ | Supplement Left Nipple with Nonautologous Tissue Substitute, Percutaneous Approach |
| --- | --- |
| 0HUX77Z | Supplement Left Nipple with Autologous Tissue Substitute, Via Natural or Artificial Opening |
| 0HUX7JZ | Supplement Left Nipple with Synthetic Substitute, Via Natural or Artificial Opening |
| 0HUX7KZ | Supplement Left Nipple with Nonautologous Tissue Substitute, Via Natural or Artificial Opening |
| 0HUX87Z | Supplement Left Nipple with Autologous Tissue Substitute, Via Natural or Artificial Opening Endoscopic |
| 0HUX8JZ | Supplement Left Nipple with Synthetic Substitute, Via Natural or Artificial Opening Endoscopic |
| 0HUX8KZ | Supplement Left Nipple with Nonautologous Tissue Substitute, Via Natural or Artificial  Opening Endoscopic |
| 0HUXX7Z | Supplement Left Nipple with Autologous Tissue Substitute, External Approach |
| 0HUXXJZ | Supplement Left Nipple with Synthetic Substitute, External Approach |
| 0HUXXKZ | Supplement Left Nipple with Nonautologous Tissue Substitute, External Approach |
| 0U5J0ZZ | Destruction of Clitoris, Open Approach |
| 0U5JXZZ | Destruction of Clitoris, External Approach |
| 0U9J00Z | Drainage of Clitoris with Drainage Device, Open Approach |
| 0U9J0ZZ | Drainage of Clitoris, Open Approach |
| 0U9JX0Z | Drainage of Clitoris with Drainage Device, External Approach |
| 0U9JXZZ | Drainage of Clitoris, External Approach |
| 0UBJ0ZX | Excision of Clitoris, Open Approach, Diagnostic |
| 0UBJ0ZZ | Excision of Clitoris, Open Approach |
| 0UBJXZX | Excision of Clitoris, External Approach, Diagnostic |
| 0UBJXZZ | Excision of Clitoris, External Approach |
| 0UCJ0ZZ | Extirpation of Matter from Clitoris, Open Approach |
| 0UCJXZZ | Extirpation of Matter from Clitoris, External Approach |
| 0UMJXZZ | Reattachment of Clitoris, External Approach |
| 0UNJ0ZZ | Release Clitoris, Open Approach |
| 0UNJXZZ | Release Clitoris, External Approach |
| 0UQG0ZZ | Repair Vagina, Open Approach |
| 0UQJ0ZZ | Repair Clitoris, Open Approach |
| 0UQJXZZ | Repair Clitoris, External Approach |
| 0UTJ0ZZ | Resection of Clitoris, Open Approach |
| 0UTJXZZ | Resection of Clitoris, External Approach |
| 0UUG07Z | Supplement Vagina with Autologous Tissue Substitute, Open Approach |
| 0UUG0JZ | Supplement Vagina with Synthetic Substitute, Open Approach |
| 0UUG0KZ | Supplement Vagina with Nonautologous Tissue Substitute, Open Approach |
| 0UUG47Z | Supplement Vagina with Autologous Tissue Substitute, Percutaneous Endoscopic  Approach |
| 0UUG4JZ | Supplement Vagina with Synthetic Substitute, Percutaneous Endoscopic Approach |
| 0UUG4KZ | Supplement Vagina with Nonautologous Tissue Substitute, Percutaneous Endoscopic  Approach |
| 0UUG77Z | Supplement Vagina with Autologous Tissue Substitute, Via Natural or Artificial Opening |
| 0UUG7JZ | Supplement Vagina with Synthetic Substitute, Via Natural or Artificial Opening |
| 0UUG7KZ | Supplement Vagina with Nonautologous Tissue Substitute, Via Natural or Artificial  Opening |
| 0UUG87Z | Supplement Vagina with Autologous Tissue Substitute, Via Natural or Artificial Opening Endoscopic |
| 0UUG8JZ | Supplement Vagina with Synthetic Substitute, Via Natural or Artificial Opening Endoscopic |
| 0UUG8KZ | Supplement Vagina with Nonautologous Tissue Substitute, Via Natural or Artificial |

|  | Opening Endoscopic |
| --- | --- |
| 0UUGX7Z | Supplement Vagina with Autologous Tissue Substitute, External Approach |
| 0UUGXJZ | Supplement Vagina with Synthetic Substitute, External Approach |
| 0UUGXKZ | Supplement Vagina with Nonautologous Tissue Substitute, External Approach |
| 0UUJ07Z | Supplement Clitoris with Autologous Tissue Substitute, Open Approach |
| 0UUJ0JZ | Supplement Clitoris with Synthetic Substitute, Open Approach |
| 0UUJ0KZ | Supplement Clitoris with Nonautologous Tissue Substitute, Open Approach |
| 0UUJX7Z | Supplement Clitoris with Autologous Tissue Substitute, External Approach |
| 0UUJXJZ | Supplement Clitoris with Synthetic Substitute, External Approach |
| 0UUJXKZ | Supplement Clitoris with Nonautologous Tissue Substitute, External Approach |
| 0VT90ZZ | Resection of Right Testis, Open Approach |
| 0VT94ZZ | Resection of Right Testis, Percutaneous Endoscopic Approach |
| 0VTB0ZZ | Resection of Left Testis, Open Approach |
| 0VTB4ZZ | Resection of Left Testis, Percutaneous Endoscopic Approach |
| 0VTC4ZZ | Resection of Bilateral Testes, Percutaneous Endoscopic Approach |
| 0VTS0ZZ | Resection of Penis, Open Approach |
| 0VTS4ZZ | Resection of Penis, Percutaneous Endoscopic Approach |
| 0VTSXZZ | Resection of Penis, External Approach |
| 0W4N071 | Creation of Penis in Female Perineum with Autologous Tissue Substitute, Open Approach |
| 0W4N0J1 | Creation of Penis in Female Perineum with Synthetic Substitute, Open Approach |
| 0W4N0K1 | Creation of Penis in Female Perineum with Nonautologous Tissue Substitute, Open Approach |
| 0W4N0Z1 | Creation of Penis in Female Perineum, Open Approach |
